# How Important Are Study Designs? A Simulation Assessment of Vaccine Effectiveness Estimation Bias with Time-Varying Vaccine Coverage, and Heterogeneous Testing and Baseline Attack Rates

**DOI:** 10.1101/2022.08.25.22279235

**Authors:** Jing Lian Suah, Naor Bar-Zeev, Maria Deloria Knoll

## Abstract

**Objective:** We studied how commonly used vaccine effectiveness (VE) study designs (variations of cohorts, and test-negative designs) perform under epidemiological nuances more prominent in the COVID-19 era, specifically time-varying vaccine coverage, and heterogeneous testing behaviour and baseline attack rates with selection on willingness to vaccinate.

**Methodology:** We simulated data from a multi-parameter conceptual model of the epidemiological environment using 888125 parameter sets. Four configurations of cohorts, and two test-negative designs, were conducted on the simulated data, from which estimation bias is computed. Finally, stratified and fixed effects linear regressions were estimated to quantify the sensitivity of estimation bias to model parameters.

**Findings:** Irrespective of study designs, dynamic vaccine coverage, and heterogeneous testing behaviour and baseline attack rates are important determinants of bias. Study design choices have non-trivial effects on VE estimation bias even if these factors are absent. The importance of these sources of bias differ across study designs.

**Conclusion:** A re-benchmarking of methodology, especially for studying COVID-19 VE, and implementation of vaccine-preventable disease surveillance systems that minimise these sources of bias, are warranted.

**Highlights:**

- This paper simulated a theoretical model with frictions in vaccination, testing, baseline disease risks, and heterogeneous vaccine effectiveness to evaluate estimation bias across four cohort and two test-negative designs.
- In theory, bias depends on behavioural asymmetries (in testing, and baseline risk) between the vax-willing and vax-unwilling, and the speed of vaccination rollout.
- There is intrinsic estimation bias across all study designs, with the direction and magnitude contingent on specific conditions.
- In scenarios that may be reflective of past SARS-CoV-2 waves, the degree of bias can be substantial, attributable to variation in assumed testing and baseline risk frictions.
- A regression-based decomposition indicates that study designs have visibly different primary sources of estimation bias, and degree of robustness in general.
- This study warrants a re-benchmarking of methodology and reporting checklists for VE research, and informs the design of cost-effective surveillance by quantifying part of the bias-implementation cost trade-off.

## 1 Introduction

When first-generation COVID-19 vaccines were rolled out via mass vaccination in early 2021, the World Health Organisation (WHO) issued an interim guidance on evaluating vaccine effectiveness (VE), based primarily on past non-COVID-19 outbreaks (WHO 2021). Reflecting this, the WHO made an explicit preference for test-negative (TND) or test-negative casecontrol (TNCC) designs in minimising confounding from health-seeking behaviour, while acknowledging the merits of retrospective and prospective cohorts. Other methods, including the screening method, and regression discontinuity design (RDD), were discussed (Orenstein et al. 1985, WHO 2021). This recommendation, and the makeup of the COVID-19 VE literature, provide the basis to evaluate variations of cohorts and TNDs.

The pre-COVID-19 literature discussed several issues pertinent to the estimation bias of VE. Firstly, under reasonable sub-100% testing sensitivities and specificities, both cohorts, and case-controls and TNDs showed small biases under simulations (Orenstein et al. 2007), but correction for misclassification bias is possible (Patel et al. 2020, Endo et al. 2020). This holds for COVID-19, given the regulations requiring high accuracy rates in professionally administered Reverse Transcription Polyamerase Chain Reaction (RT-PCR) and Rapid Antigen (RTK-Ag) tests. Secondly, TNDs are argued to be more robust to selection on health-seeking behaviour than cohorts (Jackson & Nelson 2013). Thirdly, TNDs are unbiased under a range of assumptions, but temporary non-specific immunity post-infection of similar non-target diseases, asymmetric severity between the vaccinated and unvaccinated, and, imbalances in the rates of non-target diseases in the same symptom cluster (e.g., influenza-like illnesses that are influenza and non-influenza) between the vaccinated and unvaccinated result in non-trivial biases (Foppa et al. 2013, Jackson & Nelson 2013, Jackson et al. 2018). Fourthly, study designs, when constructing counterfactuals, ought to consider different models of vaccine failure (e.g., ‘leaky’ vaccines, waning VE), confounders (e.g. time-varying exposure risk, and disease predisposition), and outcome definitions (Crowcroft & Klein 2018). The COVID-19 VE literature correctly uses specific designs to assess waning relative to static VE, and regression analysis to address confounders. Attention was also given to outcome definitions, such as differences between post-COVID-19-positive deaths and clinically audited deaths due to COVID-19, and between clinically confirmed SARS-CoV-2 infections and positive tests.

Notwithstanding the aforementioned concerns, the COVID-19 vaccine rollout in 2021-22 deviated in substantial respects from pre-pandemic vaccination distribution. Firstly, preCOVID-19 vaccine take-up tend to be stable, but not during the COVID-19 pandemic (Ritchie et al. 2020). A COVID-19-era simulation noted the importance of temporal effects arising from differential risks at equivalent post-vaccination time points (Lewnard et al. 2021). However, even with constant background risk, those vaccinated earlier have more opportunities of disease exposure post-vaccination than those vaccinated later. Secondly, high but stagnant coverage without long-run supply constraints indicate selection between the vaccinated and unvaccinated, potentially on health-seeking, testing, and risk behaviour (Ritchie et al. 2020). Thirdly, testing regimens varied, ranging from selective to comprehensive (Ritchie et al. 2020). These may present new sources of bias for COVID-19 VE studies amid vaccine rollouts.

This study asks “how do commonly used VE study designs (variations of cohorts, and TNDs) perform amid (1) time-varying vaccine coverage, (2) latent selection of and risk behaviours on willingness to vaccinate and (3) ‘leaky’ vaccines (such that *V E* < 100% against infection)?” We attempt to answer in three steps. Firstly, a theoretical discussion of estimation bias. Secondly, a simulation of the theorised environment, and VE estimates under variations of cohorts and TNDs. Thirdly, an analysis of the drivers of estimation bias.

## 2 Methods

### 2.1 Conceptual Framework and Simulation

Data are simulated from a conceptual framework with 10 adjustable parameters. There are four behavioural blocks, detailed in Appendix A — (1) vaccination take-up, (2) testing behaviour, (3) distribution of vaccine effectiveness, and (4) disease outcomes. The following parameters choices Ξ were used, amounting to 888125 unique combinations. We considered range for generalisability, and sparsity for computational ease. ‘Ideal’ conditions are when vaccine coverage is static, and behavioural frictions in testing and baseline risk do not exist (*p*_*v*_ = 1, Θ_*τ*_ = 1, *p*_*τ*_ = 1, *k*_*τ*_ = 1, *k*_*α*_ = 1).

Hence, there are four groups.

1. Unwilling to be vaccinated (vax-unwilling) and tested (test-unwilling): *θ*_*v*_ = 0 and *θ*_*τ*_ = 0
2. Unwilling to be vaccinated (vax-unwilling), but willing to be tested (test-willing) : *θ*_*v*_ = 0 and *θ*_*τ*_ = 1
3. Willing to be vaccinated (vax-willing), but unwilling to be tested (test-unwilling): *θ*_*v*_ = 1 and *θ*_*τ*_ = 0
4. Willing to be vaccinated (vax-willing) and tested (test-unwilling): *θ*_*v*_ = 1 and *θ*_*τ*_ = 1)

Theoretical bias is as follows (equations 1 to 8). *w*_*t*_ refers to the daily weights implicit to the choices of study designs, which has no closed form. Full derivation, including special cases of static vaccine coverage (*T* = 1), is in Appendix B.

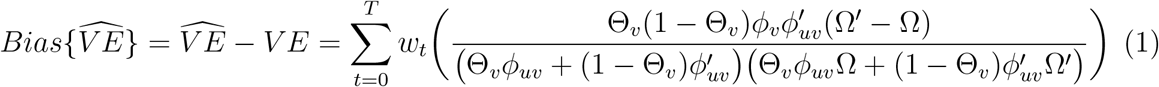

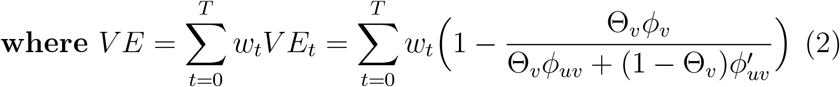

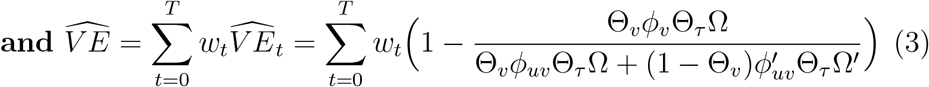

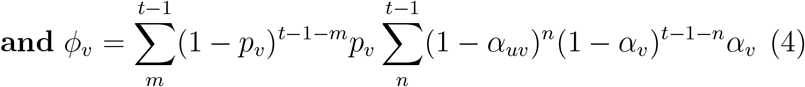

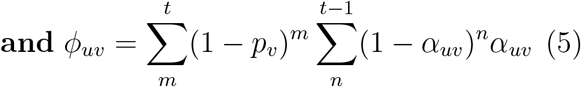

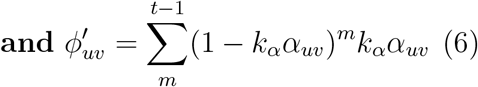

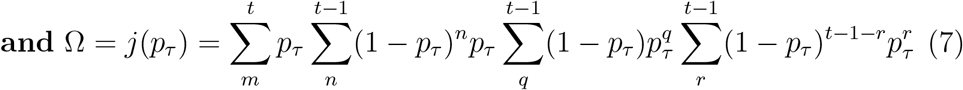

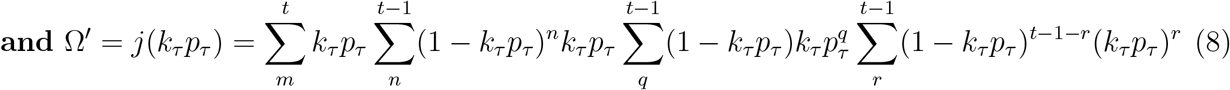

Ceteris paribus, theoretical bias decreases in *p*_*v*_ (upward bias), and increases as *k*_*τ*_ (downward) and *k*_*α*_ (upward) diverge from 1. Direction is inferred from signs in equation 1. Theoretical bias primarily depends the behavioural asymmetries between those who are vax-willing and vax-unwilling, and the speed of vaccination rollout. While the incidence of vax-willing individuals enters the equation, incidence of test-willing individuals does not, and only testing behaviour does. An unbiased design, given parameter set Ξ, optimises for *w*_*t*_, such that theoretical bias is zero.

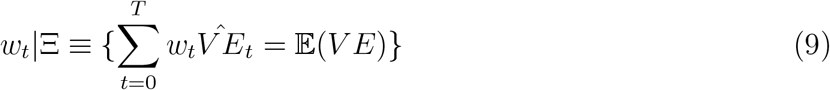

### 2.2 Study Designs

We analyse two families of study designs — (i) cohorts and (ii) test-negative designs (TNDs) — for their ubiquity and merits in the literature when handling two pertinent issues.

First, the asymmetry in testing propensity arising from *k*_*τ*_ ≠ 1. TNDs seek to address this by conditioning the study population on having being tested, hence health-seeking behaviour (Crowcroft & Klein 2018). Cohorts can control for baseline testing behaviour, e.g., number of tests taken pre-observation.

Second, time-varying vaccine coverage when *T* > 1 and/or *p*_*v*_ < 1. Unlike pre-COVID19 studies, COVID-19 vaccines are evaluated amid mass vaccination. From the conceptual framework, bias arises from that amongst the vaccinated, those vaccinated earlier have more draws of disease exposure till the end of time *T* than those vaccinated later. Ceteris paribus, breakthrough infection probability is higher for early-takers. In cohorts, researchers adjust for time-at-risk (e.g, using person-days or dynamic population as offsets in count models, using time-to-event via survival models, and controlling for timing of disease exposure or vaccination). In TNDs, the researcher may restrict to only the first positive, or first negative (if never positive) test, rather than multiple negatives per individual. These are analogous to solving for daily weights *w*_*t*_ in the conceptual model such that the probability of testing positive between time of vaccination *t*_*v*_ and the end of study *T* is independent of time.

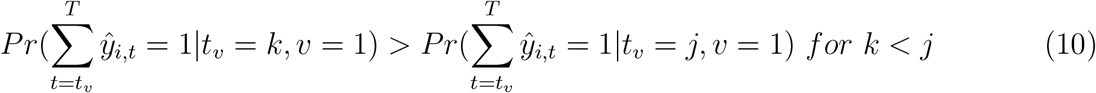

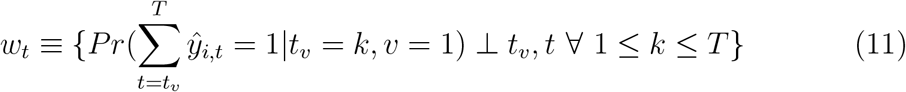

Table 2 summarises the study designs. For cohorts, we further consider when true outcomes are observable. TNDs require testing.

**Table 1:** Parameters in the Simulation

| Parameter | Meaning | Values for Simulation |
| --- | --- | --- |
| $N$ | Simulated population size | 1000 |
| $T$ | Number of chronological days used in the simulation; this is equivalent to the follow-up duration | 20, 30, 40, 50, 60 |
| $\Theta_v$ | Fraction of population that is willing to be vaccinated (vax-willing); fraction $\Theta_v$ takes values $\theta_v = 1$ and fraction $1 - \Theta_v$ takes values $\theta_v = 0$ | 0.3, 0.4, 0.5, 0.6, 0.7 |
| $p_v$ | Probability of being vaccinated if not already vaccinated but vax-willing in period $t$; $p_v = 1$ implies static coverage, such that all vax-willing start off vaccinated, and $p_v \rightarrow 0$ the opposite | 0.15, 0.3, 0.4, 0.5, 0.6, 0.85, 1 |
| $\Theta_\tau$ | Fraction of population that is willing to be tested (test-willing); fraction $\Theta_\tau$ takes values $\theta_\tau = 1$ and fraction $1 - \Theta_\tau$ takes values $\theta_\tau = 0$ | 0.5, 0.65, 0.75, 0.85, 1 |
| $p_\tau$ | Probability of being tested if test-willing in period $t$ | 0.5, 0.65, 0.75, 0.85, 1 |
| $k_\tau$ | Ratio of probability of being tested of the vax-unwilling to the vax-willing ($\frac{p_\tau^{\theta_v=0}}{p_\tau^{\theta_v=1}}$); $k_\tau < 1$ indicates that the vax-unwilling are less likely to be tested than the vax-willing | 0.5, 0.75, 0.9, 1, 1.1, 1.25, 1.5 |
| $\alpha_b$ | Baseline attack rate for the vax-willing, which is the daily probability of contracting the disease if vax-willing but unvaccinated | 0.025 |
| $k_\alpha$ | Ratio of the baseline infection rate of the vax-unwilling relative to that of the vax-willing ($\frac{\alpha_b^{\theta_v=0}}{\alpha_b}$); $k_\alpha < 1$ indicates that the baseline infection rate of the vax-unwilling is lower than that of the vax-willing | 0.5, 0.75, 0.9, 1, 1.1, 1.25, 1.5 |
| $\mu_{VE}$ | Mean of the VE distribution, $VE_i \sim \text{Beta}(\alpha = 9, \beta = \frac{\alpha}{\mu_{VE}} - \alpha)$, $0 < \mu_{VE} < 1$ for individual $i$ | 0.5, 0.6, 0.7, 0.8, 0.9 |

**Table 2:** Permutations of Study Designs

| Study Design | Approach Used to Estimate VE | Notes |
| --- | --- | --- |
| Cohort using aggregated data, with person-days as offset | Negative binomial regression (count analysis) | Commonly used where aggregate data is preferred or accessible in lieu of granular data, albeit often with a Poisson regression (Bar-On et al. 2021, Goldberg et al. 2021, Suah, Husin, Tok, Tng, Thevananthan, Low, Appannan, Zin, Zin, Yahaya et al. 2022) |
| Cohort using aggregated data, with daily population as offset | Negative binomial regression (count analysis) | Same as above but uses directly population, which may be time-varying, instead of person-days |
| Cohort using individual-level data, adjusting for immortal time bias | Cox Regression (survival analysis) | Often used in prospective cohorts where time-to-event is of interest, and where the quality of exposure-censoring time variables are robustly collected (Jara et al. 2021, n.d.) |
| Cohort using individual-level data | Logistic regression (binary outcomes) | Often used where time measures are unreliable, when time-to-event not of interest, as opposed to occurrence of outcomes, or when assumptions necessary for survival models, e.g., proportional hazards, fail (Suah, Husin, Tok, Tng, Thevananthan, Low, Appannan, Zin, Zin, Yahaya et al. 2022, Suah et al. 2021) |
| TND using individual-level data, keeping only the first positive, and, if never positive, the first negative test | Logistic regression (binary outcomes) | Often used when testing data from ambulatory care settings, healthcare facility screening, administrative registers, and ILI/SARI surveillance are deployed (Olson et al. 2022) |
| TND using individual-level data, keeping only the first positive, but allowing for multiple negative tests | Logistic regression (binary outcomes) | Used when multiple follow-up points per unique individuals are available, such as through community infection surveys (Bernal et al. 2021) |

**Table 3:**
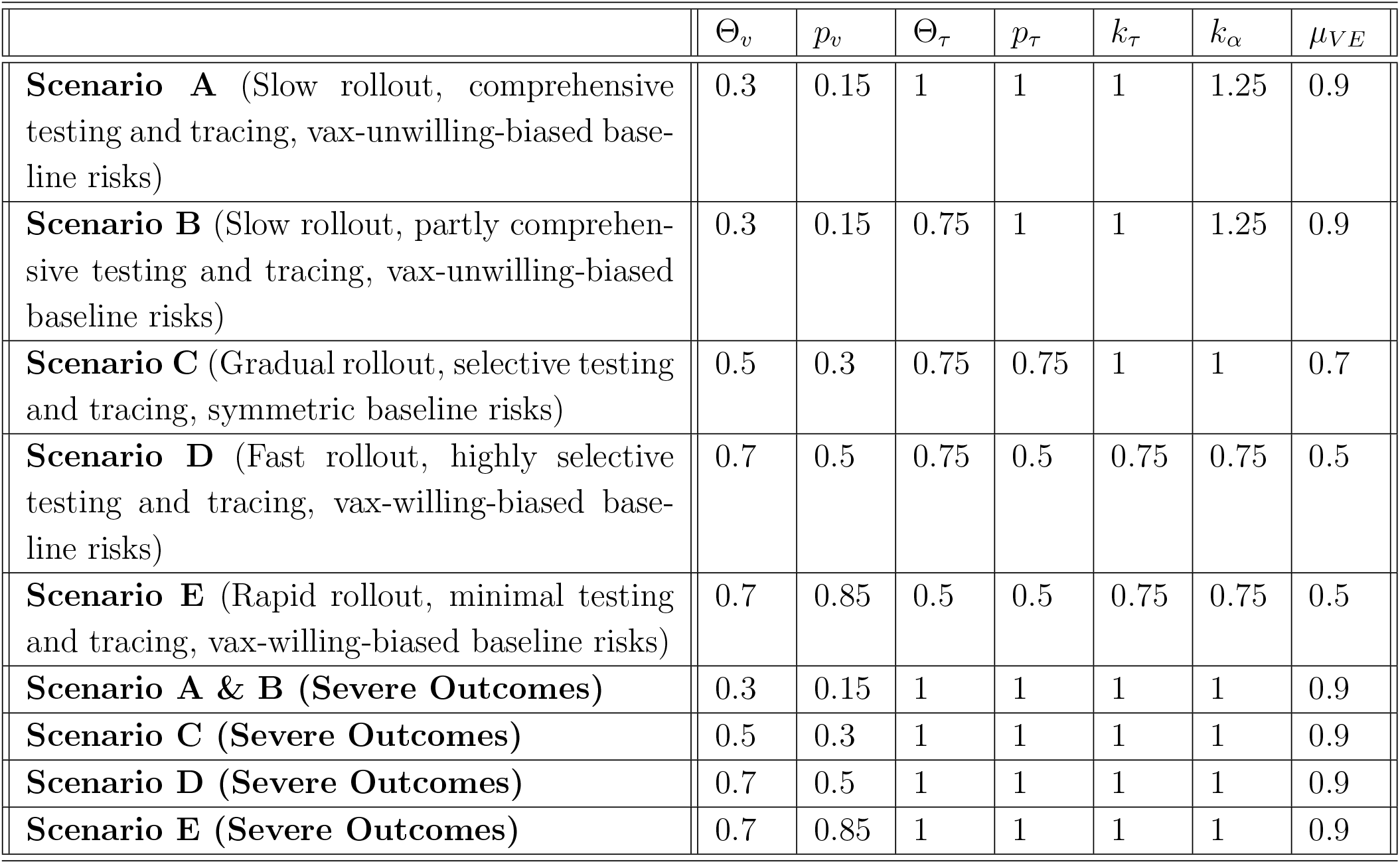
Scenario-Specific Parameter Assumptions

| | $\Theta_v$ | $p_v$ | $\Theta_\tau$ | $p_\tau$ | $k_\tau$ | $k_\alpha$ | $\mu_{VE}$ |
| --- | --- | --- | --- | --- | --- | --- | --- |
| **Scenario A** (Slow rollout, comprehensive testing and tracing, vax-unwilling-biased baseline risks) | 0.3 | 0.15 | 1 | 1 | 1 | 1.25 | 0.9 |
| **Scenario B** (Slow rollout, partly comprehensive testing and tracing, vax-unwilling-biased baseline risks) | 0.3 | 0.15 | 0.75 | 1 | 1 | 1.25 | 0.9 |
| **Scenario C** (Gradual rollout, selective testing and tracing, symmetric baseline risks) | 0.5 | 0.3 | 0.75 | 0.75 | 1 | 1 | 0.7 |
| **Scenario D** (Fast rollout, highly selective testing and tracing, vax-willing-biased baseline risks) | 0.7 | 0.5 | 0.75 | 0.5 | 0.75 | 0.75 | 0.5 |
| **Scenario E** (Rapid rollout, minimal testing and tracing, vax-willing-biased baseline risks) | 0.7 | 0.85 | 0.5 | 0.5 | 0.75 | 0.75 | 0.5 |
| **Scenario A & B (Severe Outcomes)** | 0.3 | 0.15 | 1 | 1 | 1 | 1 | 0.9 |
| **Scenario C (Severe Outcomes)** | 0.5 | 0.3 | 1 | 1 | 1 | 1 | 0.9 |
| **Scenario D (Severe Outcomes)** | 0.7 | 0.5 | 1 | 1 | 1 | 1 | 0.9 |
| **Scenario E (Severe Outcomes)** | 0.7 | 0.85 | 1 | 1 | 1 | 1 | 0.9 |

**Table 4:** Checklist for VE Research to Gauge Estimation Bias

| No. | Item | Purpose |
| --- | --- | --- |
| 1 | Pace of vaccination (daily time series or average) **during** the study period, including trend breaks, e.g., surge in doses given | To gauge $p_v$ |
| 2 | Vaccine coverage **by the end of** the study period | To gauge $\Theta_v$ (and potentially $k_v$) |
| 3 | The pace of vaccination rollout **after** the study period | To gauge $\Theta_v$ (and potentially $k_v$) |
| 4 | Testing rate in the study population, and the overall population | To gauge $\Theta_\tau$ and $p_\tau$ |
| 5 | Testing rate by vaccination status or relevant comparator groups | To gauge $k_\tau$ |
| 6 | Contact tracing regime (if relevant for disease studied) in place during the study period, e.g., comprehensive forward and backward, symptomatic forward only, no contact tracing | To gauge $\Theta_\tau$, and $p_\tau$ |
| 7 | Testing regime in place during the study period, including scope, eligibility, barriers of accessibility (direct and indirect), and selection criteria related to vaccination, e.g., universal (including asymptomatic) testing with private cost, symptomatic testing with private cost, symptomatic testing with price or queue discrimination on vaccination status | To gauge $\Theta_\tau$, $p_\tau$, and $k_\tau$ |
| 8 | Presence of NPIs to influence vaccine take-up via restrictions on disease transmission-relevant activities | To gauge $k_\alpha$ |
| 9 | **[For TNDs]** Baseline characteristics of study participants stratified by vaccination status (usually reported by test-positive, and test-negative) | To gauge $k_\alpha$, and $k_\tau$ |
| 10 | **[Where available]** VE estimates from meta-analyses, and systematic reviews specific to the disease, pathogenic variant, and clinical outcome of interest | To form priors on $\mu_{VE}$ beyond the study |

For all parameter sets Ξ, the simulated data are used to estimate the VE, *β* is the relative risk (negative binomial regression), hazard ratio (Cox regression), or odds ratio (logistic regression). Bias is the difference between estimated 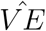 and mean of the VE distribution *μ*_*V E*_. Absolute bias 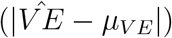 is also used.

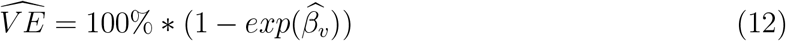

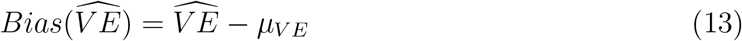

### 2.3 Scenario-Specific Parameter Assumptions

To illustrate estimation bias in practice, we isolate parameter sets that reflect various historical SARS-CoV-2 waves (A: Wild Type; B: Alpha / Beta / Gamma; C: Delta; D: Omicron, BA1-2; E: Omicron, BA3-5), such as the supply and demand of vaccines (*p*_*v*_), contact tracing and testing practices (Θ_*τ*_, *p*_*τ*_, *k*_*τ*_), and behavioural context of those unwilling to be vaccinated (*k*_*α*_). Detailed justifications are in the supplementary appendix. As surveillance of severe outcomes (hospitalisation, ICU admission, and death) is often stricter due to more specific outcome definitions, and clinical audits, these corresponding scenarios assume frictionless testing (Θ_*τ*_ = 1, *p*_*τ*_ = 1, *k*_*τ*_ = 1) and symmetric baseline risk behaviour (*k*_*α*_ = 1).

### 2.4 Drivers of Bias

To assess the relative importance of the specific parameters on simulated bias, we estimated the sensitivity of estimation bias to parameter values using two linear regressions with the ordinary least squares (OLS) estimator. Firstly, by study designs. Secondly, the average ‘within-design’ sensitivities using dummy fixed effects (FE). Technical aspects are in Appendix C. As the FE estimator returns a weighted average measure of association, divergent signs between designs are masked.

To ease interpretation, variable parameters are transformed as follows. To limit influence from outliers, estimation biases outside [−60, 60] are excluded, leaving behind 886413 parameter sets. Invariable parameters (*N*, and *α*_*b*_) are omitted.

- *k*_*τ*_ → |*k*_*τ*_ − 1|; absolute deviation from 1 (deviation from independent testing rates)
- *p*_*v*_ → |*p*_*v*_ − 1|; absolute shortfall from 1 (deviation from static vaccine coverage)
- *p*_*τ*_ → |*p*_*τ*_ − 1|; absolute shortfall from 1 (deviation from perfect testing if test-willing)
- Θ_*τ*_ → |Θ_*τ*_ − 1|; absolute shortfall from 1 (deviation from full test-willing population)
- Θ_*v*_ → |Θ_*v*_ − 1|; absolute shortfall from 1 (deviation from full vax-willing population)
- *μ*_*V E*_ → |*μ*_*V E*_ − 1|; absolute shortfall from 1 (deviation from ‘perfect’ vaccine)

## 3 Findings

The simulation took 415516 seconds (115.42 hours) in Python 3.10 on a AWS EC2 c6i.32xLarge Amazon Linux instance with 128 virtual central processing units, one of the most powerful public-accessible compute-optimised virtual machines when the analysis was conducted.

The colour range in all heat maps of estimation biases is constrained to [−20, 20]. A 20 percentage point (ppt) over-estimation puts the estimated COVID-19 VE against death from a meta-analysis of 8 platforms spanning 5 variants of concern (Higdon et al. 2022) from the neighbourhood of 60% - 99% to 40% - 79%. The equivalent for severe disease from 55% - 99% to 35% - 79%, and that of SARS-CoV-2 infection to below 50%. This hypothetical 20 ppt over-estimation represents a substantial gap in estimated, and ‘true’ protection offered. A contour map of number needed to vaccinate (NNV) and VE, given any input of the baseline attack rate *α*_0_, is also available for inspection in the supplementary appendix. For sufficiently low baseline attack rates (< 1%), as is likely the case for severe COVID-19, a 20 ppt change in VE represents sizeable change in NNV, even if VE is high. E.g., changing VE from 90% to 70% when the baseline attack rate is 1% raises the NNV from 100 to 150, a 50% increase in NNV. The colour scheme differentiating over-(red) and under-estimation (blue) reflects that over-estimation 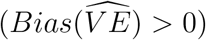 may be more damaging, as policymakers may be overconfident in unwinding public health restrictions.

### 3.1 Simulation

The simulated performance of each design differs across parameter sets Ξ (full table of 888125 parameter combinations is here in supplementary materials; a snippet is in the appendix). Under ‘ideal’ conditions (*p*_*v*_ = 1, Θ_*τ*_ = 1, *p*_*τ*_ = 1, *k*_*τ*_ = 1, *k*_*α*_ = 1; figure 1), some designs are equivalent. Under perfect testing, cohorts where tests are required to observe disease outcomes and where tests are not required are identical. If testing is comprehensive, the TND with single test contribution is identical to the binary outcomes cohort. In the ‘ideal’ case, the TND that allows for multiple negative tests is the least biased, followed closely by the survival analysis cohort. The count analysis cohort with population as the offset underestimates, while the binary outcomes cohort, and TND with single test contribution overestimates. As follow-up lengthens (*T*) under ‘ideal’ conditions, estimation biases of the binary outcome cohort, single test TND, and count analysis cohort with population as the offset increase.

**Figure 1:**
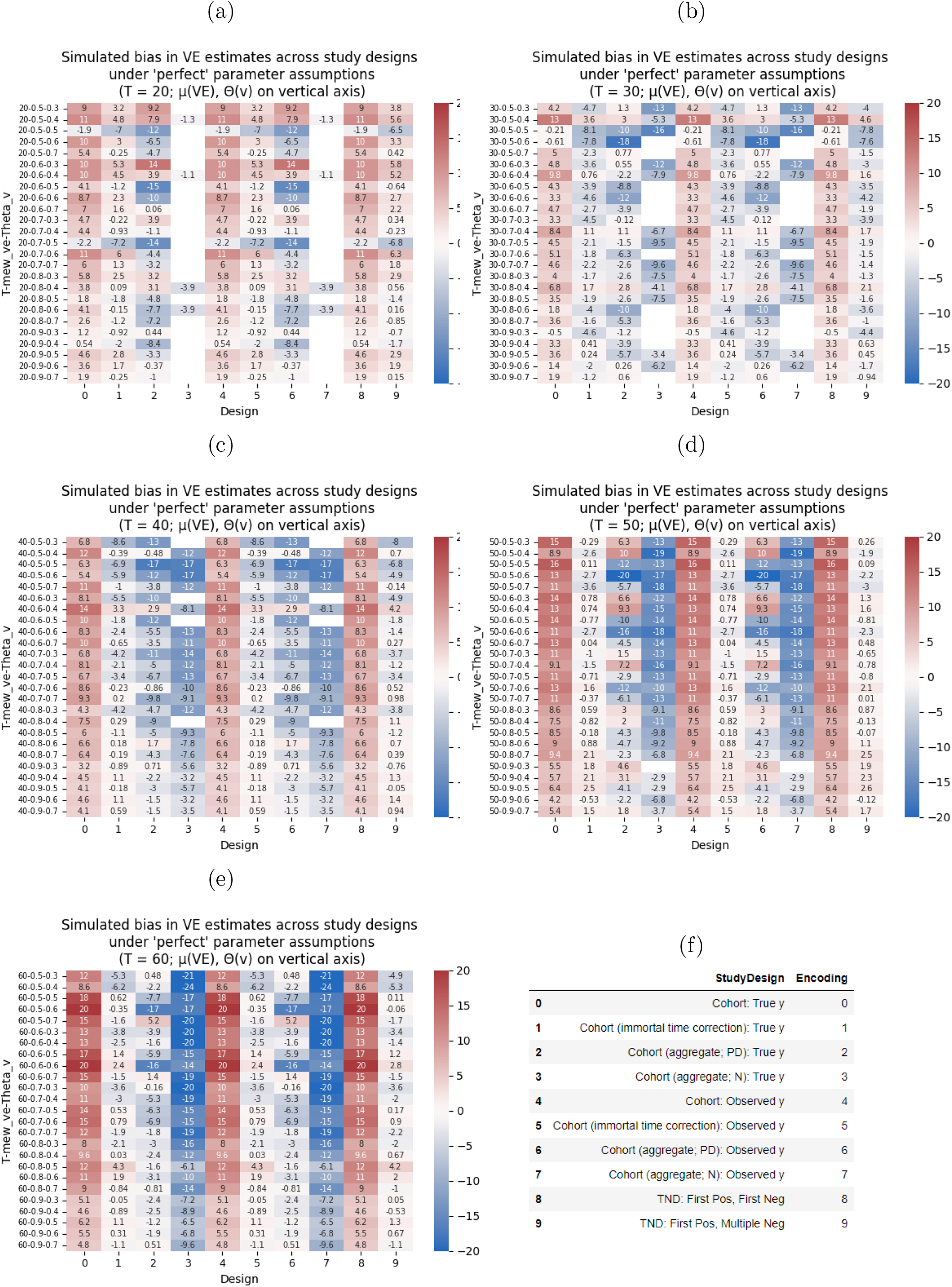
Simulated estimation bias under ‘ideal’ parameter assumptions

Under gradual rollout (*p*_*v*_ ≠ 1, all else ideal), the opposite holds. Relaxing instead perfect testing (Θ_*τ*_ < 1 and/or *p*_*τ*_ and/or *k*_*τ*_ ≠ 1, all else ideal), simulated biases change in a mixed manner. Lowering only the population share of test-willing Θ_*τ*_ reverses some findings. Next, some relative performances are reversed if pre-completion of rollout, and perfect testing are relaxed (*p*_*v*_ < 1, Θ_*τ*_ < 1, *p*_*τ*_ < 1, *k*_*τ*_ ≠ 1). Asymmetry in baseline attack rates between the vax-willing and vax-unwilling (*k*_*α*_ ≠ 1) tend to increase the degree of bias. Higher baseline attack rates for the vax-unwilling relative to the vax-willing tend to overstate VE, while the opposite tend to understate. Without long-run supply constraints, everyone who is willing to be vaccinated will be vaccinated. If long-run vaccine coverage is stable but not 100%, the vaccinated and unvaccinated may be fundamentally different in behaviour. Hence, studies comparing the two groups under high vaccine coverage may be biased.

The estimation biases vary under all nine scenarios-specific parameter sets (figure 2). Count analysis cohorts were generally biased downwards. In scenario A where parameters were least deviated from ‘ideal’, biases were low. In scenarios B and C, survival analysis cohorts, and TNDs with multiple negative tests performed consistently better. In scenario C, binary outcomes cohort were comparable to the former designs, but with an upward bias. In scenarios D and E, the TND with multiple negative tests was marginally least biased, and downward bias is sizeable across all designs. As frictionless testing, and symmetric baseline risk (Θ_*τ*_ = 1, *p*_*τ*_ = 1, *k*_*τ*_ = 1, *k*_*α*_ = 1) are assumed in the analogues of severe outcomes, the only source of bias is the speed of vaccination rollout (*p*_*v*_). Hence, estimation bias is minimal except for the count analysis cohorts. For completeness, the supplementary appendix formally analyses the stability of relative ranks between pairwise designs.

**Figure 2:**
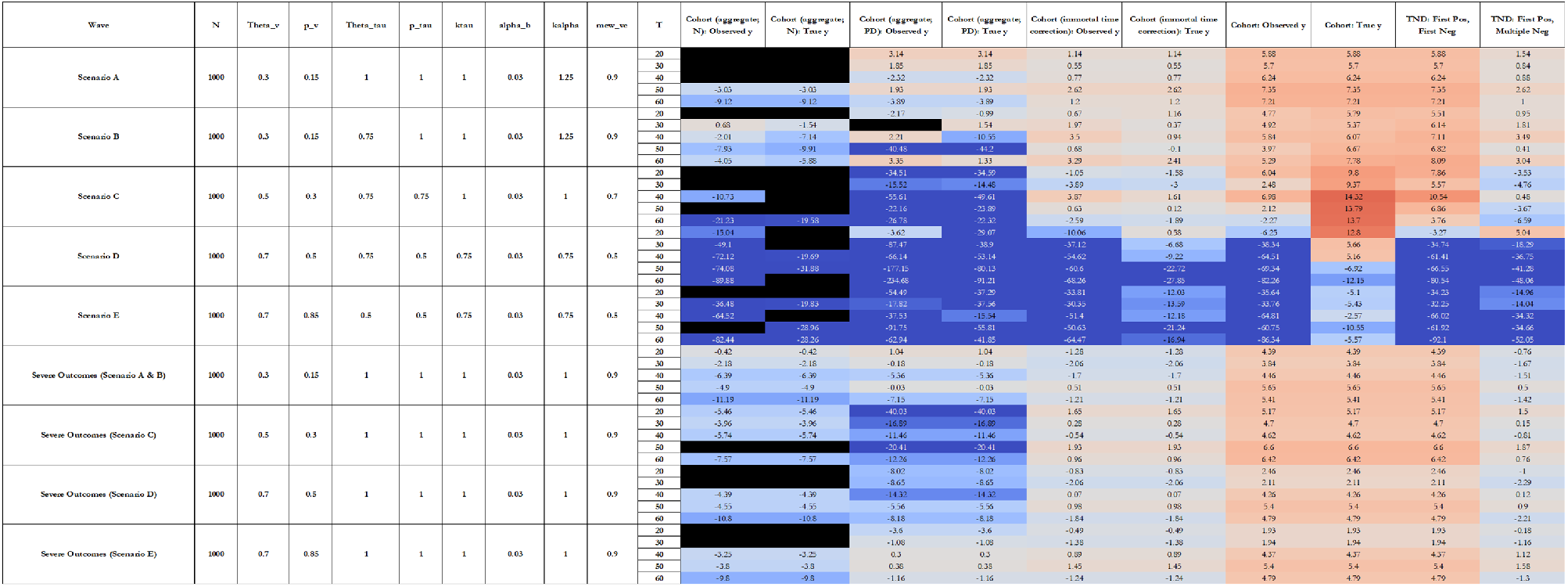
Simulated estimation bias under scenario-specific parameter sets

### 3.2 Drivers of Bias

Figure 3 summarises the sensitivity of estimation bias to parameters by study design. Figure 4 shows the average sensitivity, and the parameter with the largest sensitivity. The appendix contains a version with sensitivities rescaled to scenario D (*p*_*v*_ = 0.5; Θ_*τ*_ = 0.75; *p*_*τ*_ = 0.5; *k*_*τ*_ = 0.75; *k*_*α*_ = 0.75), and another version that reflects scenario D with steps taken to mitigate testing frictions (*p*_*v*_ = 0.5; Θ_*τ*_ = 0.75; *p*_*τ*_ = 0.75; *k*_*τ*_ = 0.95; *k*_*α*_ = 0.75) to gauge potential gains for policymakers if surveillance mitigate selective testing, and the FE model estimates as a summative exercise. This ‘rescaling’ assists researchers who often lack choice on methodology, a direct result of pre-existing surveillance and data structures, gauge the estimation bias they are working with.

**Figure 3:**
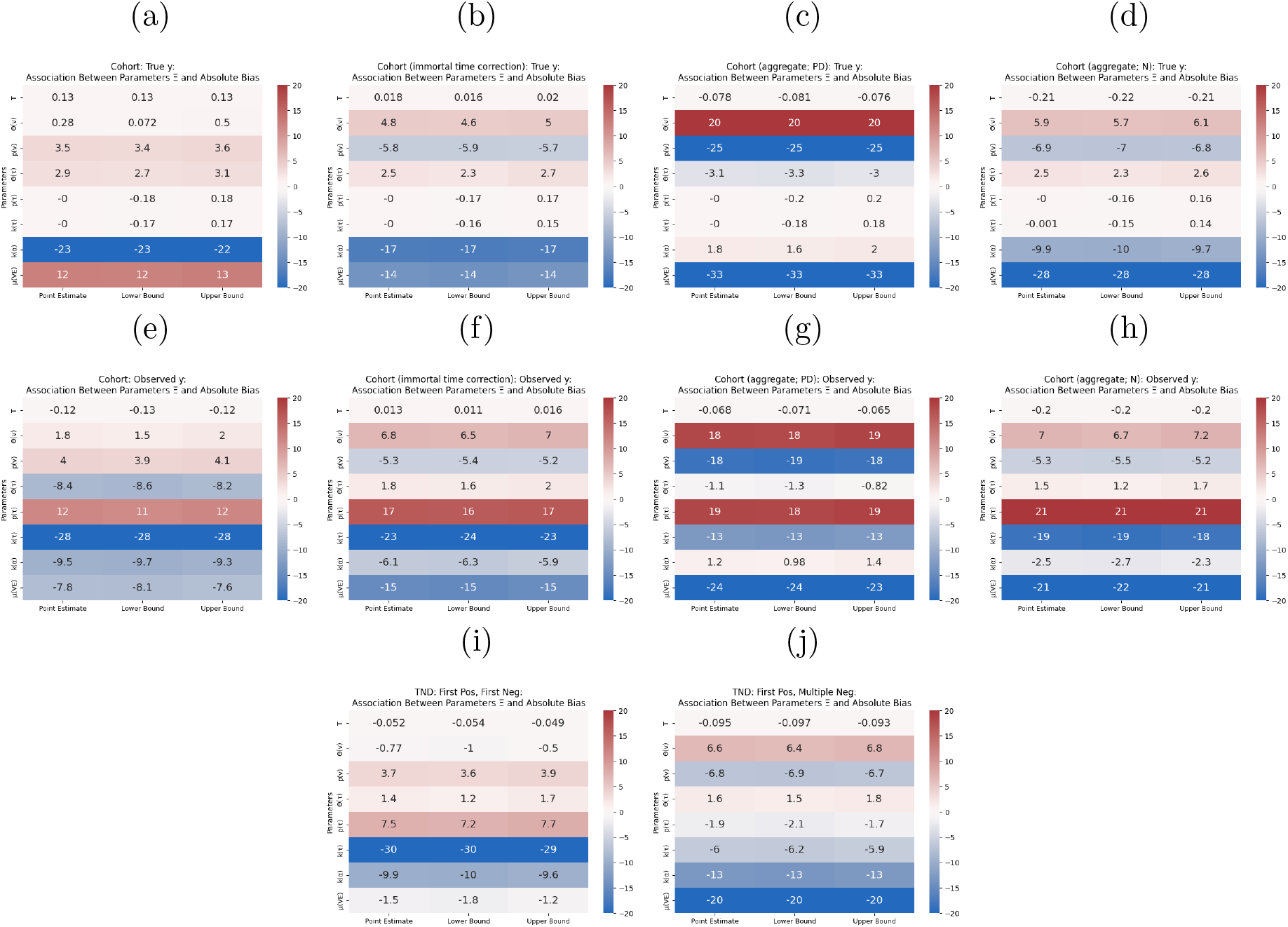
Association Between Parameters Ξ and Absolute Bias By Study Design

**Figure 4:**
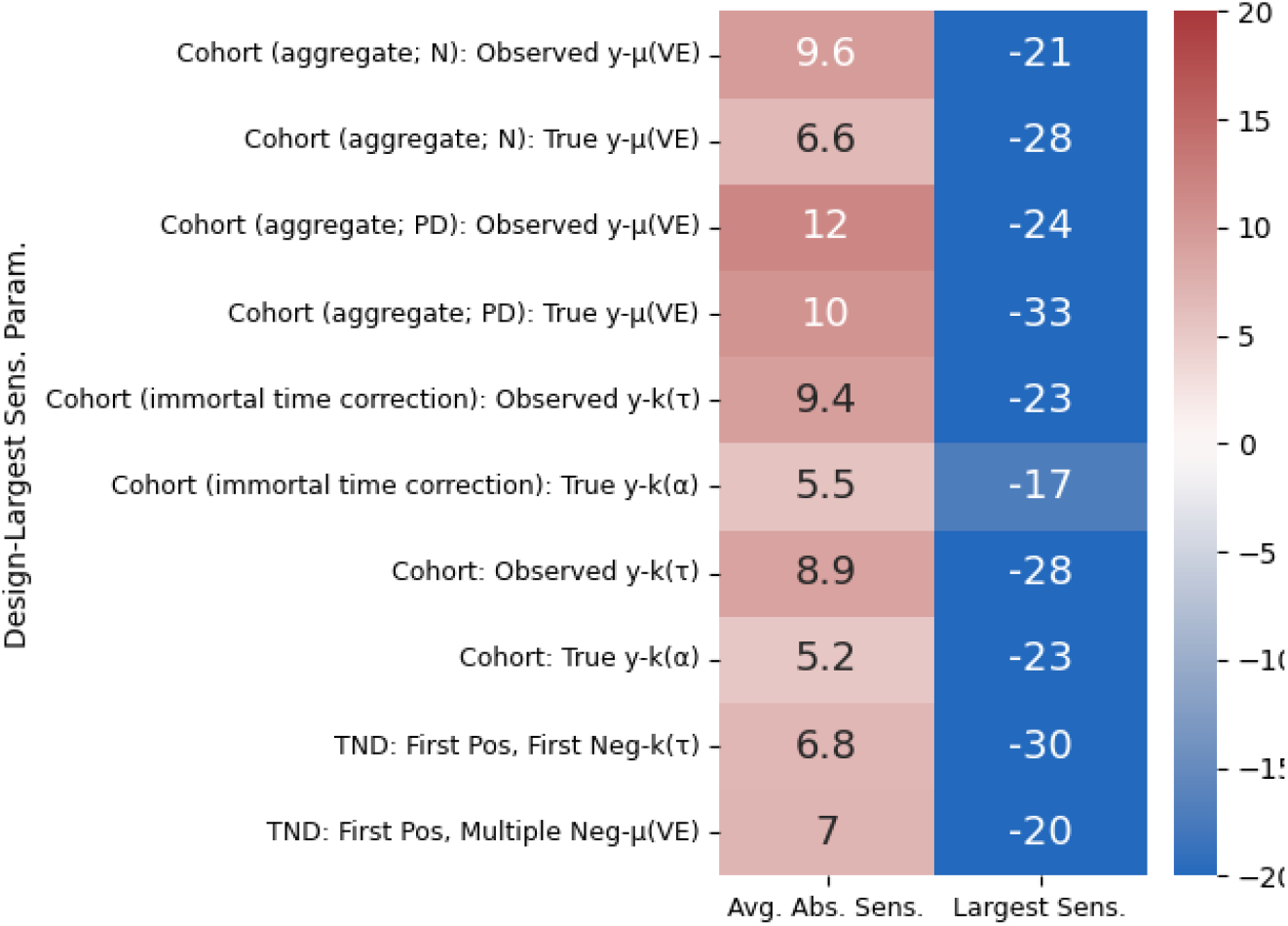
Average Absolute and Largest Sensitivity Between Parameters Ξ and Absolute Bias (Parameter With Largest Sensitivity on Vertical Axis)

Across all designs except the binary outcomes cohort without testing requisites, deviation in the mean of the VE distribution (*μ*_*V E*_) from ‘perfect’ protection (100%) exerts a downward bias. This reflects asymmetry in the beta distribution, as all estimators (MLE on negative binomial, logistic, and Cox regressions) require log-normal errors, which could be unrealistic in reality.

When true outcomes are observable (figures 3a to 3d), in binary outcomes and survival analysis cohorts, the asymmetry in baseline attack rates between the vax-willing and vax-unwilling (*k*_*α*_) is important. In the latter, deviation from static vaccine coverage (vaccination rate, *p*_*v*_) is salient. In count analysis, cohorts with person-days as offset, the deviation from static vaccine coverage (vaccination rate, *p*_*v*_; downward bias) is most important, but secondary to the asymmetry in baseline attack rates between the vax-willing and vax-unwilling (*k*_*α*_) in the case with population as offset.

When testing is prerequisite to observe outcomes (figures 3e to 3h), in binary outcomes and survival analysis cohorts, the asymmetry in testing propensity between the vax-willing and vax-unwilling (*k*_*τ*_), and the deviation from perfect testing amongst the test-willing population (*p*_*τ*_) are most important. Of secondary importance are the asymmetry in baseline attack rates between the vax-willing and vax-unwilling (*k*_*α*_), the deviation from static vaccine coverage (vaccination rate, *p*_*v*_), full test-willing population (Θ_*τ*_), and full vax-willing population (Θ_*v*_). In count analysis cohorts, the asymmetry in testing propensity between the vax-willing and vax-unwilling (*k*_*τ*_), the deviation from static vaccine coverage (vaccination rate, *p*_*v*_), perfect testing amongst the test-willing population (*p*_*τ*_) and full vax-willing population (Θ_*v*_) are most important.

In TNDs with only one test per individual (figure 3i), only the asymmetry in testing propensity between the vax-willing and vax-unwilling (*k*_*τ*_) stood out, followed by the deviation from perfect testing amongst the test-willing population (*p*_*τ*_), and the asymmetry in baseline attack rates between the vax-willing and vax-unwilling (*k*_*α*_). As this configuration aims to remove the influence of timing of vaccination on the risk of disease exposure, the deviation from static vaccine coverage (vaccination rate, *p*_*v*_) has a small association with bias, similarly for deviation from full test-willing population (Θ_*τ*_). In contrast, asymmetry in baseline attack rates between the vax-willing and vax-unwilling (*k*_*α*_) is important in the TND with multiple negative tests per individual (figure 3j), followed by the asymmetry in testing propensity between the vax-willing and vax-unwilling (*k*_*τ*_), and the deviation from full vax-willing population (Θ_*v*_), and static vaccine coverage (vaccination rate, *p*_*v*_).

Finally, the average absolute sensitivity between parameters and absolute bias indicate that, amongst study designs where testing is prerequisite to observing outcomes, both specifications of TNDs are most robust to ‘less-than-ideal’ conditions. These properties make the TND attractive under dynamic vaccine rollout, and imperfect testing. The largest sensitivity measure is roughly equal across all designs.

## 4 Discussion

Where vaccine rollouts are gradual, testing is heterogeneous, and baseline risk associated with vaccination attitudes, study design choices have non-trivial effects on VE estimation bias. This extends a pre-COVID-19 review that noted study design may be important in uncovering specific models of vaccine failures and underlying mechanisms (Crowcroft & Klein 2018). Our research is unique in that it formalises, without loss of generality, the interaction between study design and the epidemiological environment in affecting estimation bias.

We modelled directly the epidemiological nuances of COVID-19 vaccines governing the pace of rollout, preference heterogeneity in testing, risk behaviour, and vaccination, and uncertainty in unobservable individual-level VE. Open access and official data globally indicate these properties (Ritchie et al. 2020). Pre-COVID-19, vaccine coverage remained stable over years, such as rotavirus vaccines (Hungerford et al. 2018). Where coverage is stable and high, such as BCG, Pol3, DTP3, and IPV (Vanderslott et al. 2013), those unwilling to be vaccinated may be systematically different from those willing. Coverage of these vaccines are similar to COVID-19 vaccines. Balanced take-up, such as PCV3, YFV, and Rotavirus vaccines (Vanderslott et al. 2013), may indicate supply constraints, such that selection in baseline risk and testing on vaccination preference may be less prominent.

Our study simulated data from a multi-parameter conceptual model of the epidemiological environment, reflective of when COVID-19 vaccines were rolled out. Hence, we can pin down the effects of (1) parameters, and (2) study designs on estimation bias. Suppose realworld data is used, both will be mixed with multidirectional selection bias. Moreover, a straightforward decomposition of bias allows researchers to understand the design-specific parameter-bias nexus. The analysis is generalisable to the increasing number of studies on effectiveness relative to vaccinated subgroups, e.g., primary vaccinated (Suah, Tng, Tok et al. 2022, Bar-On et al. 2021, Ranzani et al. 2022), and single booster vaccinated (Regev-Yochay et al. 2022). The ‘baseline’ group for behavioural frictions is simply the intended reference, rather than the unvaccinated. Going forward, the same principle applies to studies comparing next-generation vaccines, or variant-updated vaccines against existing first-generation vaccines.

Our work has three implications.

Firstly, they warrant a re-benchmarking of methodology. Recommendations for VE research, such as by the WHO (WHO 2021), need to reflect these considerations, as recent novel diseases are materially different from earlier ones during which most designs were evaluated. Our analysis of the drivers of estimation bias confirm that (1) the parameter-bias nexus differs across designs, and (2) some designs are more robust to sources of bias. In practice, researchers can use the heat maps of bias-parameter sensitivities to ‘adjust’ VE estimates, by first ascertaining the design used, and second calibrate parameters in the conceptual model, e.g., a study during the Omicron wave should have large deviations from ‘ideal’ values in the testing parameters, but small deviations in vaccination rate. Confidence intervals of the VE estimates can be expanded this way. Alternatively, researchers may adjust the point estimate and the confidence interval concurrently uni-directionally. For readers of empirical research, VE estimates may be adjusted ex post to assess where the ‘true’ VE may lie, e.g., VE estimates during the Omicron wave may require substantial upward adjustments, or wider confidence bands, than those from the Delta wave. Hence, to help form judgment on the extent of estimation bias, we recommend future research to report the following, in addition to the STrengthening the Reporting of OBservational studies in Epidemiology (STROBE) checklist.

Secondly, this study informs policymakers on designing cost-effective vaccine-preventable infectious disease surveillance systems by quantifying part of the estimation bias versus implementation cost trade-off, crucial for resource-constrained countries, or when outbreaks require temporary, cheap, surveillance. While count and survival analyses cohorts perform best under ‘ideal’ circumstances, TNDs are more resilience against sources of bias. Countries may gear surveillance in major outbreaks to use TNDs, rather than cohorts, which require more comprehensive data infrastructures. Conceptually, the policymaker ‘chooses’ a surveillance system that (1) explicitly mitigates sources of bias, and (2) enables the least biased study design, subject to a resource constraint. Both (1) and (2) are dynamically related, as some designs perform better under ‘ideal’ circumstances, but not otherwise. This motivates a formal analysis on cost-effective design optimisation. Moreover, given the importance of testing and risk behaviour heterogeneity in VE estimation bias, surveillance systems may include community random sampling, e.g., the United Kingdom’s COVID-19 Infection Survey (ONS 2022).

Thirdly, disease progression, or severity of symptoms, that is correlated with testing propensity can be discussed. In principle, this entails another layer of outcome post-infection with a unique relationship with vaccination status and testing preferences. However, this requires an excessively complex theoretical model with minimal gains in insights. Within the current model, this is best captured via testing rate (*p*_*τ*_) and asymmetry (*k*_*τ*_). Combinations of low testing rate amongst the test-willing (*p*_*τ*_ ≪ 1), and low testing propensity amongst the vax-willing than the vax-unwilling (*k*_*τ*_ ≫ 1) represent instances where symptoms are milder for the vaccinated, who are then less likely to test unless sufficiently symptomatic.

There are limitations in this study. Firstly, tests with sub-100% sensitivity and specificity is not modelled. However, a pre-COVID-19 simulation found that reasonable sensitivities and specificities (80% to 90%) yielded minimal bias (Jackson & Rothman 2015). Secondly, reinfections are not explicitly modelled. Suppose past infections provide some protective effect that compound with that of vaccines, then greater prevalence of undetected infections amongst the vaccinated than the unvaccinated may overestimate VE. An example is if breakthrough infections are extremely mild but close to each other, such that viral loads are sufficiently low to not trigger antigen tests, then estimated VE is high when true VE is low. This warrants more involved modelling in future research. Thirdly, observed confounders are not modelled explicitly. Doing so requires strong and non-generalisable assumptions on the structure between confounders, vaccination choice, and disease dynamics. Fourthly, waning VE (or building of VE) are also not modelled explicitly, which could exponentiate the complexity of the simulation. Further research may consider various forms of non-linearity in waning, interaction with behavioural frictions, and probabilistic waning.

## 5 Conclusion

Using simulated data from a multi-parameter conceptual model of the epidemiological environment, we offer three findings. Firstly, both dynamic vaccine coverage, and heterogeneous testing behaviour and baseline attack rates that are selected on willingness to vaccinate, are important determinants of VE estimation bias. Secondly, study designs have non-trivial effects on estimation bias, even without these frictions. Thirdly, these factors have different relative and absolute importance in driving estimation bias. Moving forward, a re-benchmarking of methodology, and surveillance systems of vaccine-preventable disease that minimise these sources of bias, are warranted.

## Data Availability

All scripts required for replication are available at github.com/suahjl/vemethod-simulation-frictions.

## Ethical consideration

As this research uses only simulated data, and does not involve any human participants, institutional review board (IRB) approval is not required.

## Data and replication statement

This research uses only simulated data, and does not involve any human participants. All scripts required for replication are available at **github.com/suahjl/vemethod-simulationfrictions**.

## Acknowledgement

The authors would like to thank the following individuals:

- Sheamini Sivasampu (Institute for Clinical Research, National Institutes of Health, Ministry of Health, Malaysia) for contributions in conceiving broad research questions;
- Masliyana Husin (Institute for Clinical Research, National Institutes of Health, Ministry of Health, Malaysia) for material support in securing access to AWS EC2, and administrative support; and
- Boon Hwa Tng (Research and Modelling Unit, Central Bank of Malaysia, Malaysia; contributed while on secondment at the Institute for Clinical Research, National Institutes of Health, Ministry of Health, Malaysia) for comments and suggestions related to the theoretical model, and post-simulation analysis of estimation bias.

## A Conceptual Framework (Theory)

The key assumptions of the conceptual framework are as follows.

1. Vaccination coverage at least increases over time (vaccination rollout is either pre-completed, or ongoing throughout the study)
2. A share of the population is vaccination-willing (*vax-willing* ; conversely vaccination-unwilling or *vax-unwilling*) but may not necessarily receive the vaccine
3. A share of the population is test-willing (conversely, test-unwilling) but may not necessarily test for the disease
4. The vax-unwilling’s probability of testing, suppose test-willing, may differ from that of the vax-willing
5. Vaccine effectiveness differs between individuals, but is distributed along a particular mean/mode
6. Suppose not yet infected, then the risk of infection follows a memoryless process
7. All individuals can be infected at most once, hence no re-infections
8. The baseline infection risk of the vax-unwilling may differ from that of the vax-willing Hence, there are four groups.

1. **The vax-willing, and test-willing** — will get tested, and/or vaccinated at some probability to be determined
2. **The vax-willing, but test-unwilling** — will get vaccinated at some probability, but will never get tested
3. **The vax-unwilling, but test-wiling** — will get tested at some probability, but will never get vaccinated
4. **The vax-unwilling, and test-unwilling** — will never get vaccinated, nor get tested

These assumptions allow us to zoom into the aspects highlighted in the study question — (i) time-varying vaccine coverage, latent selection of (ii) testing, and (iii) risk behaviour, specifically on attitude towards vaccination.

### A.1 Vaccination Take-Up

Over *T* time periods (e.g., days), *n*_*v,t*_ are vaccinated on day *t*. By the end of time, *t* = *T, N*_*v,t*_ are vaccinated, and *N*_*uv,t*_ are unvaccinated.

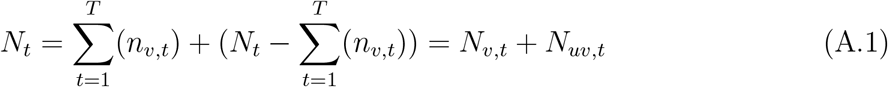

Whether an individual *i* subscribes to the vaccination drive depends on a the vax-willingness parameter (*θ*_*v,i*_). The population share of individuals who are vax-willing is Θ_*v*_. Vaxunwilling individuals never get vaccinated. This parameter is determined ex ante (before *t* = 1). Conditional on being vax-willing (*θ*_*v,i*_ = 1) and not already vaccinated, the probability of being vaccinated at period *t* is *p*_*v*_. How close *p*_*v*_ is to 1 therefore determines the pace of the rollout, with *p*_*v*_ = 1 corresponding to a static coverage throughout *t* = 1 till *t* = *T*.

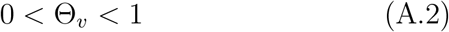

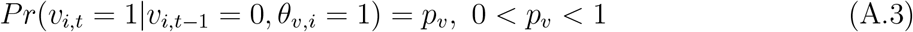

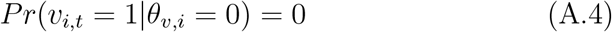

### A.2 Testing Behaviour

A fraction Θ_*τ*_ of the population is willing to undergo testing (test-willing). Test-willing individuals are indicated by their test-willingness parameter *θ*_*τ*_ = 1 (test-unwilling take *θ*_*τ*_ = 0). Individual *i*, suppose test-willing, will undergo testing at period *t* at probability *p*_*τ*_. This process is, similarly, memoryless. Test-unwilling individuals never get tested.

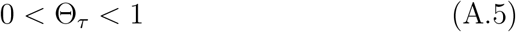

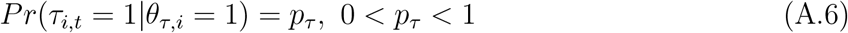

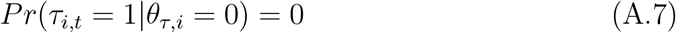

However, the probability of testing amongst vax-unwilling but test-willing individuals (*θ*_*v,i*_ = 0 and *θ*_*τ*_ = 1) is a fraction *k*_*τ*_ ≤ 1 that of the vax-willing and test-willing (*θ*_*v,i*_ = 1 and *θ*_*τ*_ = 1)

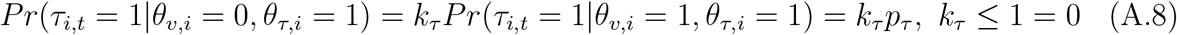

### A.3 Distribution of Vaccine Effectiveness

The motivation of modelling VE as a random variable stems from immunogenic studies suggest that both humoral (S-antibody, and B-cell) and cellular (T-cell) responses against SARS-CoV-2 differ between individuals. While no formal correlates of protection against SARS-CoV-2 have been established, it is a reasonable assumption that VE is dynamic. From an estimation perspective, statistical methods then recover the central measure (often mean) of VE in a study population.

The VE for individual *i* is a random variable that follows a Beta distribution with a mean of *μ*_*V E*_, bounded by 0 and 1, and with the *α* parameter preset to 9. A Beta distribution has two desirable properties — (1) bounded distribution, as, in theory, non-inferior vaccines have intrinsic *V E* ∈ [0%, 100%], and (2) smooth but variable slopes (a strong contender is the Triangular distribution, which is smooth, bounded, but has invariant slopes). The choice of *α* = 9 is arbitrary, and chosen for its effect on the concentration around the mean and mode; other researchers can choose higher *α* values for a more ‘concentrated’ distribution, and lower alpha values for the opposite. *μ*_*V E*_ = 0.5 (VE of 50%) corresponds to a special case where the mode of the VE distribution equates its mean. Individual *i*, if vaccinated, will experience an attack rate *α*_*v,i*_ that is lower than if unvaccinated *α*_*uv*_ by the drawn VE. This constructs individual-specific counterfactuals. For simplicity, the attack rate for the unvaccinated *α*_*uv*_ is a scalar, i.e., constant for everyone that is unvaccinated.

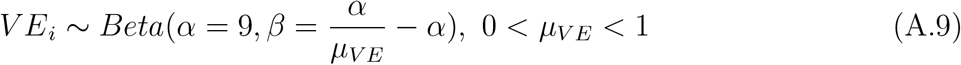

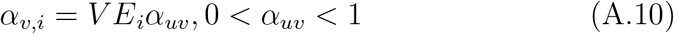

### A.4 Disease Outcome

The infection risk of individual *i* (*α*_*i*_) is a function of vax-willingness *θ*_*v,i*_, vaccination status *v*_*i,t*_, and the latent individual-specific vaccine effectiveness *V E*_*i*_, which activates if a vaccine is administered. *α*_*base*_ refers to the baseline risk for vax-willing individuals (*θ*_*v,i*_ = 1), and is scaled by *k*_*α*_ for the vax-unwilling (*θ*_*v,i*_ = 0). If vaccinated (*v*_*i*_ = 1), then the infection risk for individual *i* is further scaled by the effectiveness of the vaccine 1 − *V E*_*i*_. *k*_*α*_ is therefore the ratio of baseline infection risk of the vax-unwilling to that of the vax-willing. Moreover, we assume no re-infections. If not yet infected, the risk of infection (attack rate) for individual *i* is memoryless.

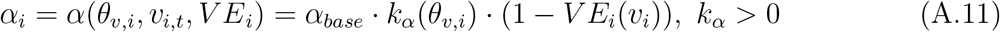

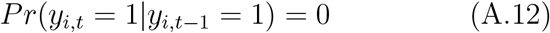

Hence, there are three permutations of infection risks, depending on vax-willingness (*θ*_*v,i*_) vaccination status (*v*_*i,t*_). (1) *α*_*vw*′,*uv*_ = *k*_*α*_*α*_*uv*_ for the vax-unwilling, and unvaccinated, (2) *α*_*vw,uv*_ = *α*_*uv*_ for the vax-willing, but unvaccinated, and (3) *α*_*vw,v*_ = *α*_*v*_ for the vax-willing, and vaccinated. The vax-unwilling, and vaccinated combination is paradoxical, hence does not exist, *α*_*vw, v*_ = *k*_*α*_*α*_*v*_ = 0.

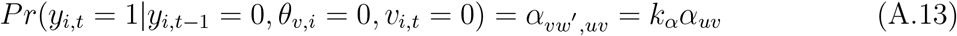

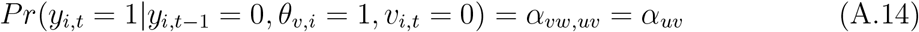

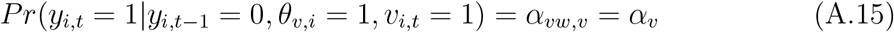

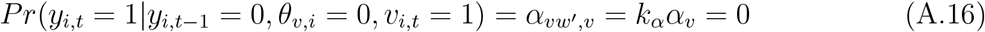

To observe infection *ŷ* at period *t*, individual *i* needs to be tested at the period of infection *τ*_*i,t*_ = 1. We can generalise to a longer detection period, but this is unnecessary convolution for the aims of the study. Tests are assumed to be perfectly sensitive and specific in this framework.

Hence, if an individual *i* is infected at period *t* but does not take a test, the observed state will not indicate infection *ŷ*_*i,t*_ = 0. If an individual is indeed not infected at period *t*, taking a test will indicate a lack of infection *ŷ*_*i,t*_ = 0. In contrast, taking a test at period *t* when infected at period *t* will observe an infection *ŷ*_*i,t*_ = 1.

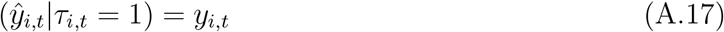

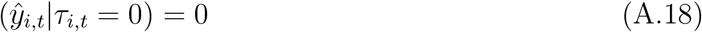

Therefore, absent of a universally test-willing population with a 100% testing rate, observed infection 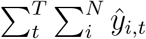 is likely a fraction of true infection 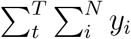.

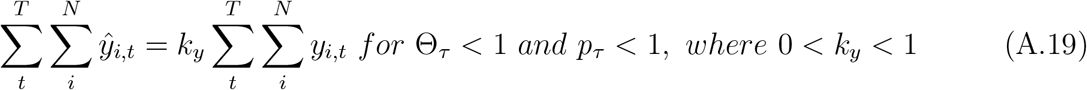

## B Theoretical Vaccine Effectiveness and Bias

VE is a function of the attack rate amongst the vaccinated relative the unvaccinated. Empirically, this is calculated from estimates of hazard ratios (survival analysis), odds ratios (binary outcomes), and relative rates (count analysis). Hence, bias is the difference between observed and true (calculated from true infection *y*) VE.

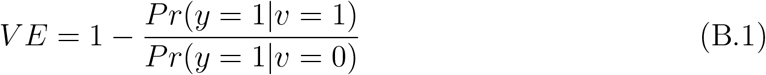

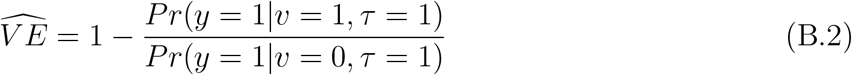

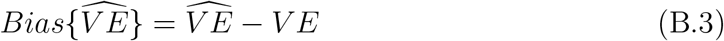

We explore the theoretical bias of VE as the following assumptions are incrementally introduced — (1) differences in baseline attack rate between the vax-willing, and vax-unwilling, (2) differences in test-willingness, and selection in testing on vax-willingness, as well as (3) gradual rollout of vaccines.

### B.1 Static: Selection in Risk on Vax-Willingness

Drawing from our conceptual model, the unvaccinated group is comprised of individuals who are willing to be vaccinated, but not vaccinated (e.g., due to supply constraints), as well as individuals who are unwilling to be vaccinated, whose baseline attack rates differ. Unbiased VE is recovered by comparing between the vaccinated and unvaccinated, conditional on being vax-willing. This is analogous to randomising the vaccinated and unvaccinated arms, such that there is no self-selection into either arms based on willingness to be vaccinated. Absent of this adjustment, the unvaccinated group is the average of the vax-willing and vax-unwilling, weighted by population share, whose baseline attack rates may differ due to fundamental differences in risk behaviour. Hence, a vanilla comparison of the vaccinated and unvaccinated arms in the presence of differential risk behaviour between the vax-willing and vax-unwilling results in biased VE estimates.

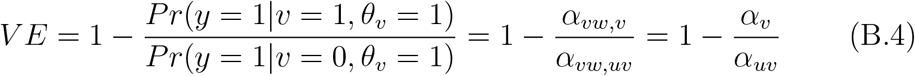

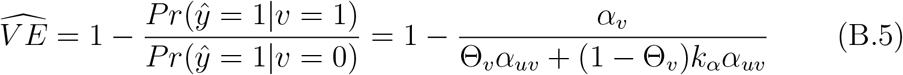

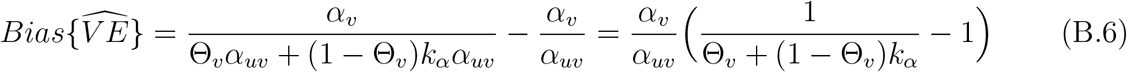

### B.2 Static: Selection in Risk and Testing on Vax-Willingness

For simplicity, we begin with the special case where there is only one time period (*T* = 1), but adding heterogeneity in testing rates based on vax-willingness. True VE is hence a function of the population share of vax-willing individuals, the probability of being vaccinated, and attack rates. The observed VE then further depends on the population share of test-willing, and the probability of being tested, as well as the relative testing propensity of the vax-unwilling to the vax-willing. Due to the asymmetry in the testing propensity and baseline attack rates between the vax-willing and vax-unwilling, the bias of observed VE is not 0. Bias is only zero if both testing propensity and baseline attack rates are independent of vax-willingness.

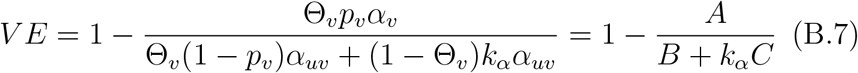

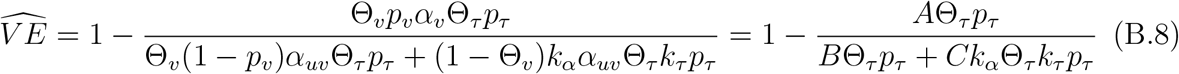

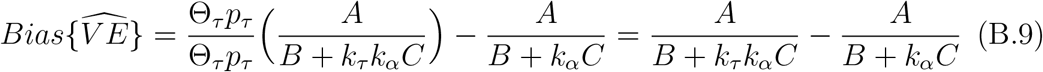

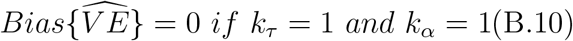

### B.3 Dynamic

In the dynamic case, there is more than one period (*T* > 1). We can think of overall *V E* as a weighted sum of period-specific *V E*_*t*_. Weights depend on the *t*-specific share of the subgroups. The specific functional form is not necessary to analyse bias in observed overall 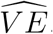.

Period-specific *V E*_*t*_, which is expressed in terms of actual outcome *y*, is a function of vaccination rate (probability of receiving the vaccine, if vax-willing and not already vaccinated) *p*_*v*_, population share of vax-willing individuals Θ_*v*_, the baseline attack rate *α*_*uv*_, and the ratio of the baseline attack rate amongst the vax-unwilling relative to the vax-willing *k*_*α*_.

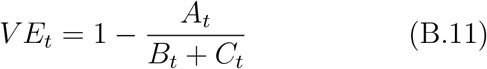

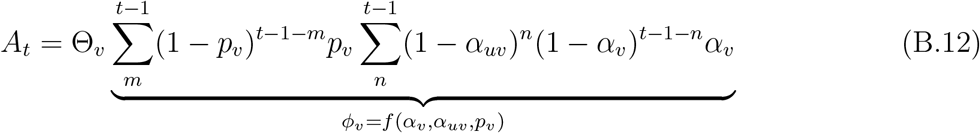

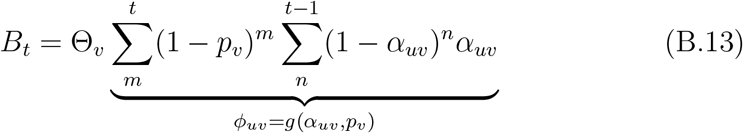

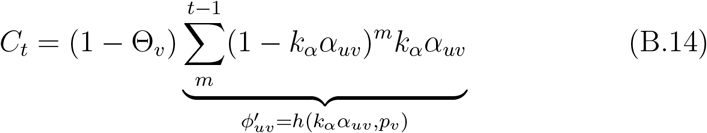

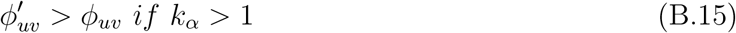

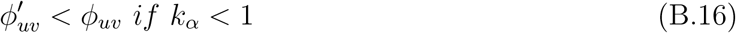

Observed period-specific 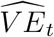 depends on the rate of testing (probability of testing if test-willing) *p*_*τ*_, population share of test-willing individuals Θ_*τ*_, and the ratio of testing probability amongst the vax-unwilling relative to the vax-willing, suppose test-willing *k*_*τ*_.

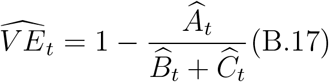

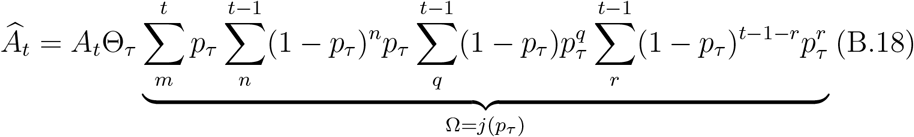

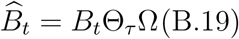

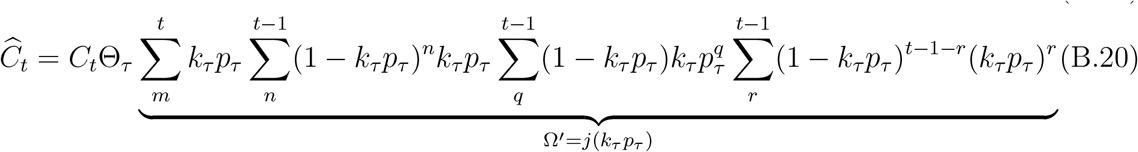

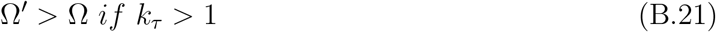

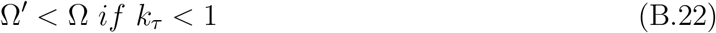

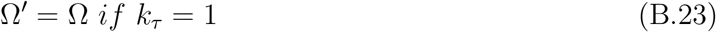

The bias of overall VE is then the difference between the weighted sums of the daily observed VEs and that of the daily true VEs.

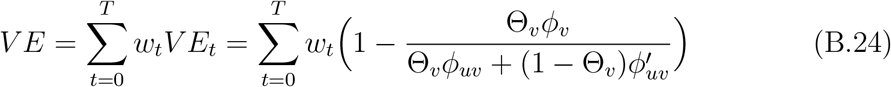

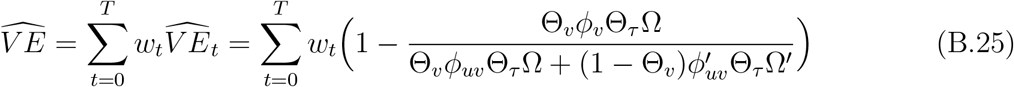

In this framework, the relative propensity of testing between the vax-unwilling and the vax-willing determines if observed 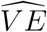 is unbiased. Independence between testing rate and vax-willingness (*k*_*τ*_ = 1) yields zero theoretical bias 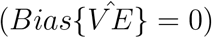.

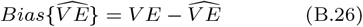

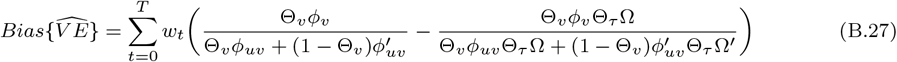

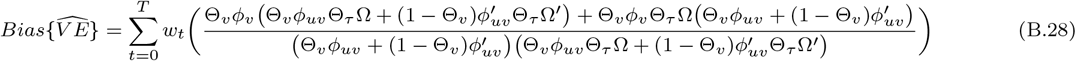

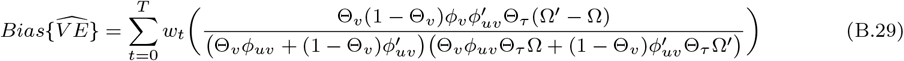

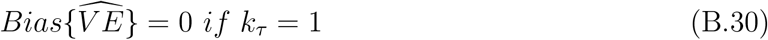

All else held equal, theoretical bias is increasing in *T* (longer time period) but decreasing in *p*_*v*_ (probability of being vaccinated if vax-willing and not already vaccinated, which both *ϕ*_*v*_ and *ϕ*_*uv*_ are functions of). Theoretical bias also increases as *k*_*τ*_ (asymmetry in testing propensity between the vax-willing and vax-unwilling) and *k*_*α*_ (asymmetry in baseline attack rates between the vax-willing and vax-unwilling) diverge from 1.

*w*_*t*_ can be interpreted as the functional form of the choice of study design. All other parameters Ξ held equal, an unbiased study design solves for *w*_*t*_, such that the dynamic *V E* equation is exactly the expectation of the *V E* distribution. The choice of *w*_*t*_ may result in unbiased estimates of *V E* for particular sets of Ξ but not necessarily for some. An unbiased design with *w*_*t*_, given parameters Ξ, further requires the observed 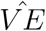 equation to yield the expectation of the *V E* distribution.

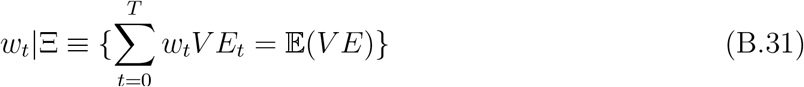

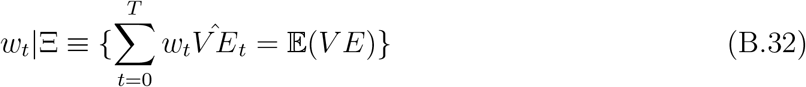

## C Scenario-Specific Parameter Assumptions

Scenario A entails slow vaccine rollout due to supply constraints arising from nascent manufacturing chains, that is limited to mostly trial settings. Comprehensive contact tracing and testing regimes were more common; together with a generally risk averse environment, imperfections in testing parameters are set to be limited. The vax-unwilling are further assumed to have higher baseline attack rates than the vax-willing due to disease-naivety and that this group is likely a selective category of disease deniers, e.g., that of COVID-19. This most likely reflects realities during the initial SARS-CoV-2 wave (Wild Type).

Scenario B assumes then that production of vaccines were scaled up, and testing became less comprehensive due to containment fatigue in the population, as well as stressed healthcare resources due to higher case load. This is more reflective of the end of 2020, when the Alpha, Beta, and Gamma variants became dominant.

Scenario C assumes that vaccines being rolled out at a faster pace, as major pharmaceutical companies were beginning to achieve economies of scale in production and orders. Due to even more extensive case load, testing became even patchier. Baseline attack rates are independent of attitudes towards vaccination. The Delta wave, which started in the second quarter of 2021, where lockdowns were imposed in response to Delta’s perceived severity relative to earlier variants, making baseline risks symmetric between the vax-willing and vax-unwilling.

Scenario D assumes that vaccine production achieved economies of scale, and orders saturated, hence vaccine rollouts were rapid. Testing became more selective, comprehensive contact tracing and testing regimes are gradually abandoned. Baseline attack rates were also lower for the vax-unwilling than the vax-willing. This is reflective of the first Omicron wave at the end of 2021, contact tracing, testing, and self-regulatory behaviour on risky social activities were gradually abandoned due to lower perceived severity of Omicron SARS-CoV-2 infections amid a high speed vaccine rollouts. A substantial fraction of countries were rolling out booster doses at this point. The vax-willing during this period may have a higher baseline risk due to a combination of repeated and undetected past infections amongst the unvaccinated, and vaccine mandates in place conditioning higher risk social activities on vaccination.

Scenario E further assumes that testing is even patchier as all forms of contact tracing, and mandated testing are abandoned. Moreover, vaccination rate is generally stable, as production lines have likely been optimised for efficiency. In the second Omicron wave at the start of 2022 as by this point, most countries have already completed their vaccination drives. Demand, rather than supply, is the constraint.

## D Drivers of Bias

Let *δ*_*M*,Ξ_ be the set of simulated bias for parameters Ξ, and design choice *M*. Further let *δ*_Ξ_|*M* be the set of bias for the specific design *M*, and Ξ_*M*_ be the parameter set for that choice of design. We then express the vector of combined simulated biases *δ*_*M*,Ξ_ as a linear combination of design fixed effects *λ*_*M*_, parameters Ξ, and a design-parameter-specific error term *ε*_*M*,Ξ_. We may allow the bias-parameter nexus to vary across design choices, in lieu of a linear fixed effects term. Hence, we express the bias conditional on design *M, δ*_Ξ_|*M* to be a linear combination of parameter set Ξ_*M*_, and an error term *ε*_Ξ|*M*_.

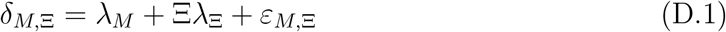

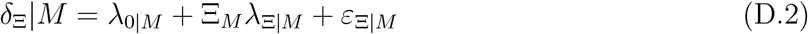

## E Additional Findings

### E.1 Rational for Range of Colours in Heat Maps

The following charts show the contour maps of (1) the number needed to vaccinate (NNV) against VE, and (2) the first derivative of NNV against VE, both conditional on baseline attack rates between 1% and 11% at 2 ppt intervals. The algebraic derivation is as follows, where *α*_*b*_ is the baseline attack rate, and *α*_*v*_ the vaccinated attack rate.

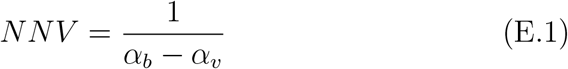

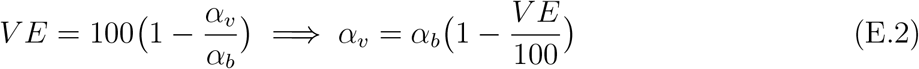

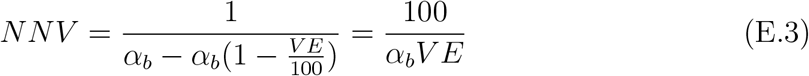

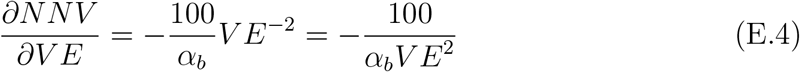

There are three points to make here. Firstly, for any VE, NNV rises at an increasing rate as the baseline attack rate (*α*_*b*_) decreases. Secondly, irrespective of *α*_*b*_, NNV falls at a decreasing rate as VE rises. Thirdly, as *α*_*b*_ decreases, the rate of decline in NNV with respect to VE becomes more pronounced.

**Figure E.1:**
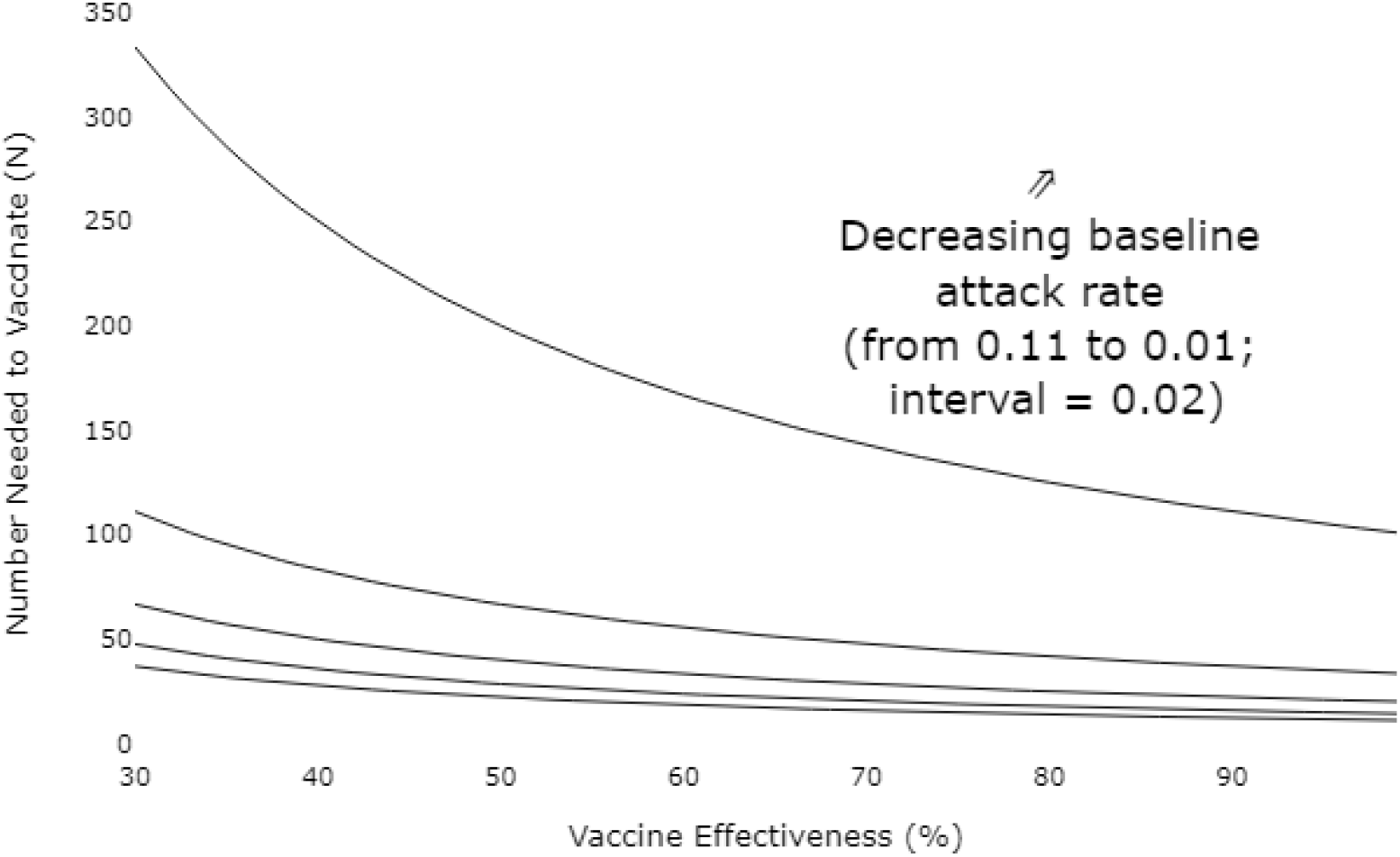
Contour Map of the Number Needed to Vaccinate (NNV) Against Vaccine Effectiveness (VE), Conditional on Baseline Attack Rates

### E.2 Simulation

The following table contains a snippet of the full table of simulated bias across study designs for all 888125 parameter sets Ξ. The full document can be found here.

**Figure E.2:**
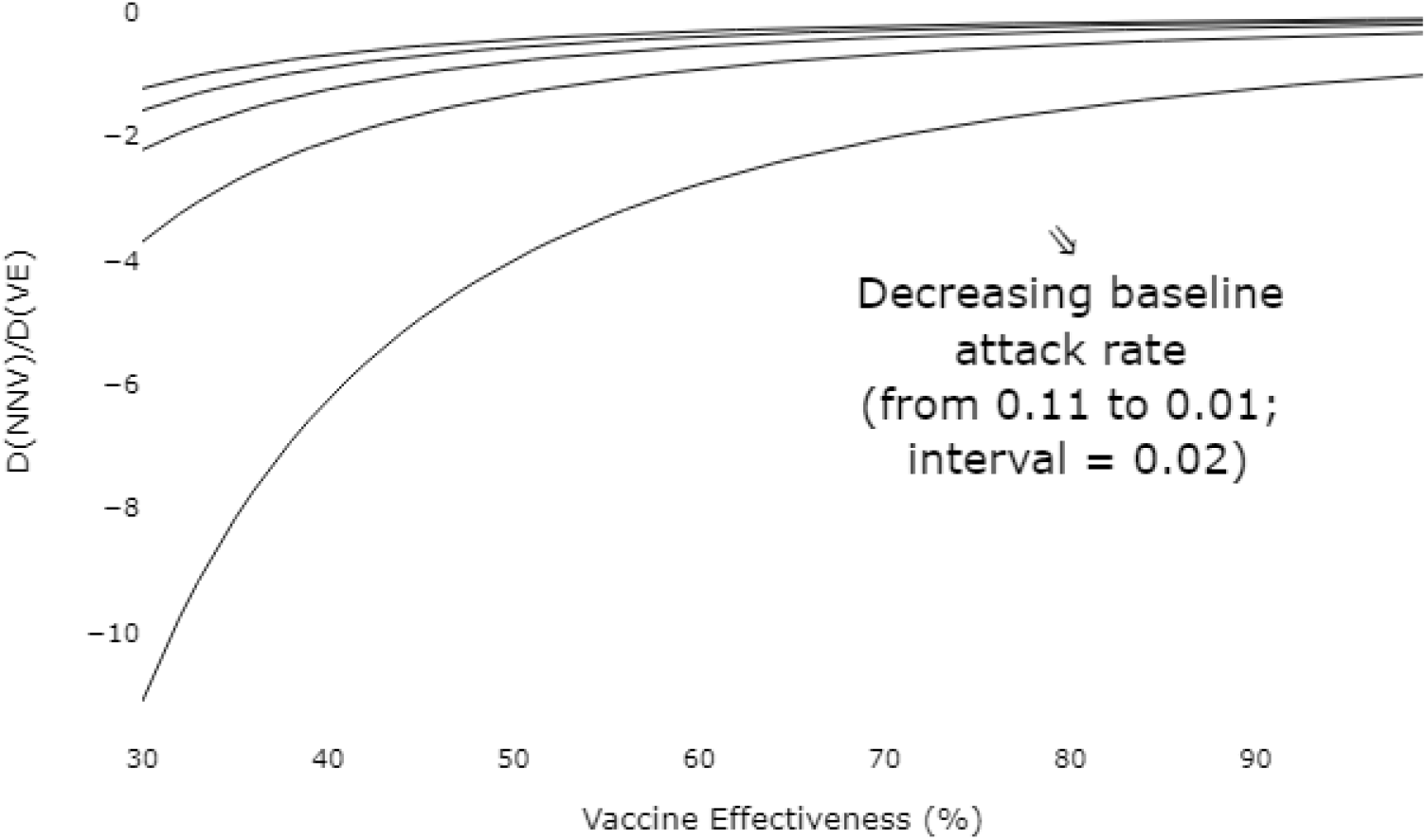
Contour Map of the First Derivative of the Number Needed to Vaccinate (NNV) Against Vaccine Effectiveness (VE), Conditional on Baseline Attack Rates

**Figure E.3:**
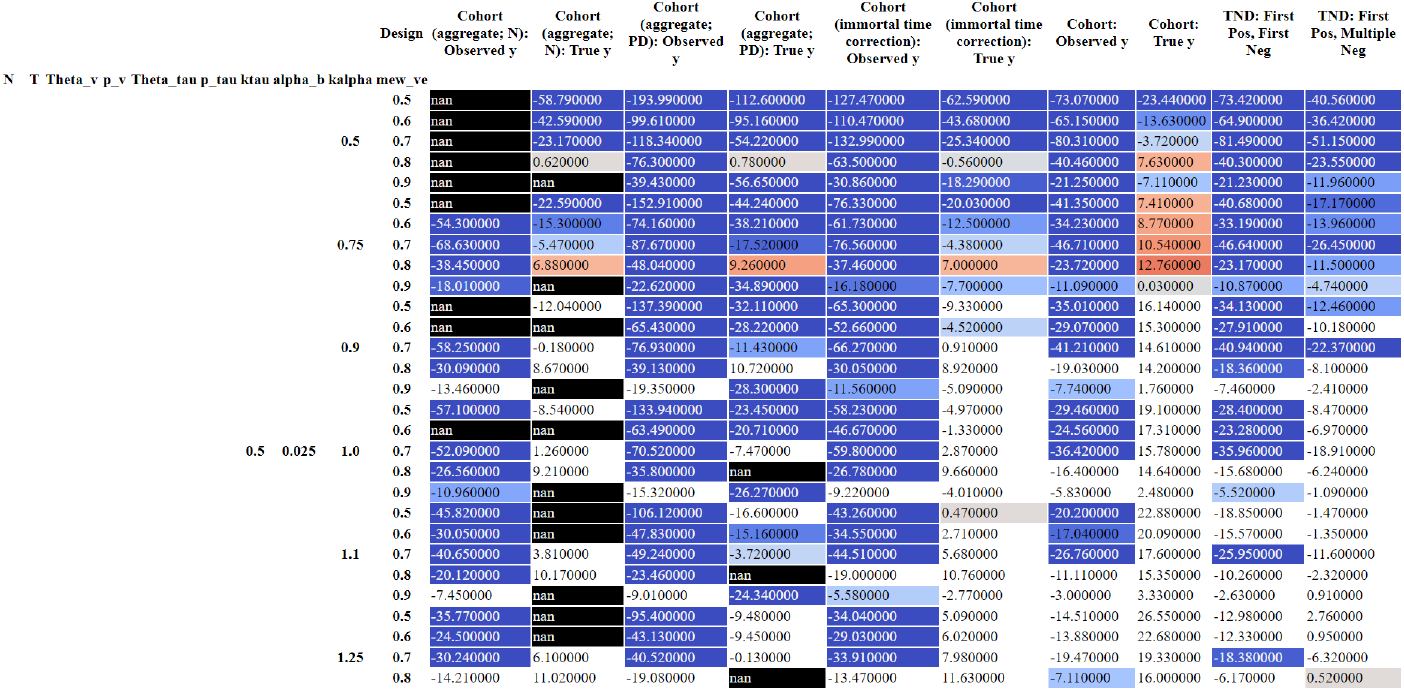
Simulated bias in VE estimates across study designs *\*Image shows a segment of the full figure, which is available as a separate file with 888125 rows*

An alternate version below normalises the simulated biases row-wise (by parameter sets), such that they are directly benchmarked against the survival analysis cohort without testing requisites. The full document can be found here.

**Figure E.4:**
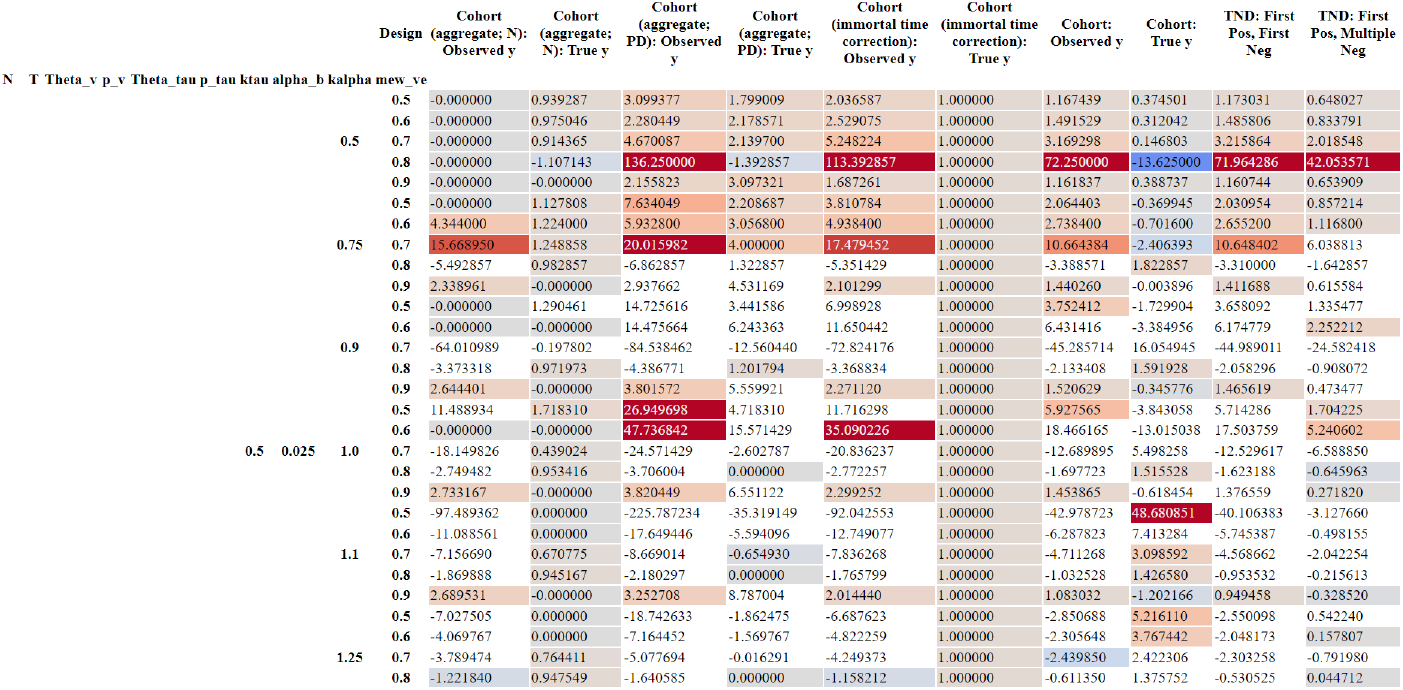
Relative bias in VE estimates across study designs; normalised to the survival analysis cohort without testing requisites *\*Image shows a segment of the full figure, which is available as a separate file with 888125 rows*

The researcher may instead be interested in a direct comparison of the study designs in terms of absolute bias. The version below first converts all estimation biases into absolutes, and then normalise to that of the survival analysis cohort without testing requisites. The full document can be found here.

**Figure E.5:**
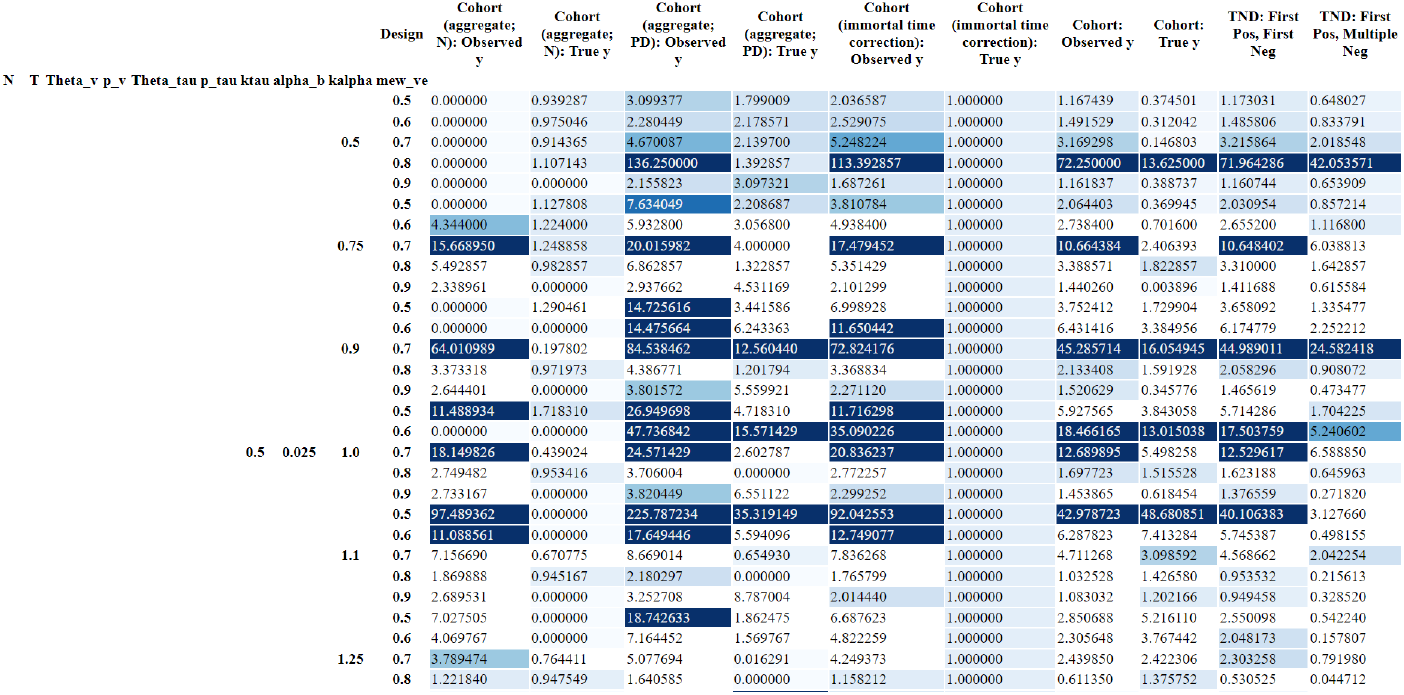
Relative absolute bias in VE estimates across study designs; normalised to the survival analysis cohort without testing requisites *\*Image shows a segment of the full figure, which is available as a separate file with 888125 rows*

In the following figure, subfigure (a) shows the % of parameter sets for which one study design yields higher VE than another, vice versa. Subfigure (b) shows the equivalent in number of parameter sets. This pairwise comparison of all 10 study designs across parameter sets suggest that rank reversals are common. However, even though there is no study design that maintained pairwise superiority for more than 80% of parameter sets, these rankings do not account for the ‘realism’ or real-world frequency of the corresponding parameter settings. Hence, when weighted against the real-world occurrence of respective parameter sets (e.g., settings reflective of Omicron as in scenarios D and E may be more likely than others), rank reversals may be infrequent, and that dominant study designs may exist.

**Figure E.6:**
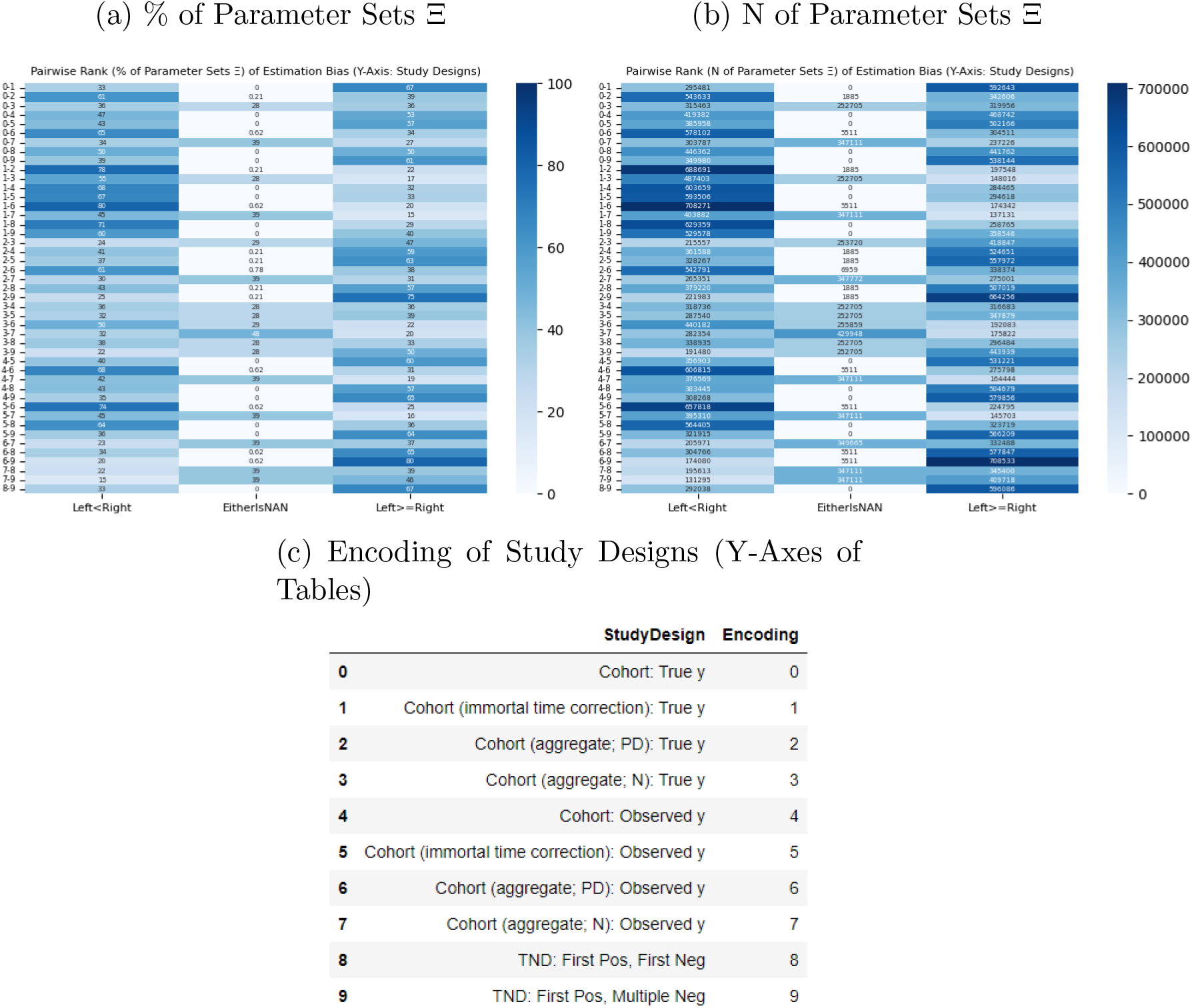
Pairwise Rank of Estimation Bias (Y-Axis: Study Designs)

### E.3 Drivers of Bias

When rescaled to parameter values in scenario D, bias is less severe in most study designs, except those highly affected by dynamic vaccine coverage (*p*_*v*_), and testing imperfections (Θ_*τ*_, *p*_*τ*_, *k*_*τ*_). Reflective of parameter choice (*k*_*α*_ = 0.75), deviation from symmetric baseline attack rates between the vax-willing and vax-unwilling may bias VE estimates only by a small but visible degree. The testing rate amongst the test-willing (*p*_*τ*_), asymmetry in testing rates between the vax-willing and vax-unwilling (*k*_*τ*_), and vaccination rate (*p*_*v*_) still exert meaningful bias on VE estimates. Importantly, as these sensitivity estimates are homothetic, the TNDs remain most robust to ‘less-than-ideal’ conditions than the cohorts.

**Figure E.7:**
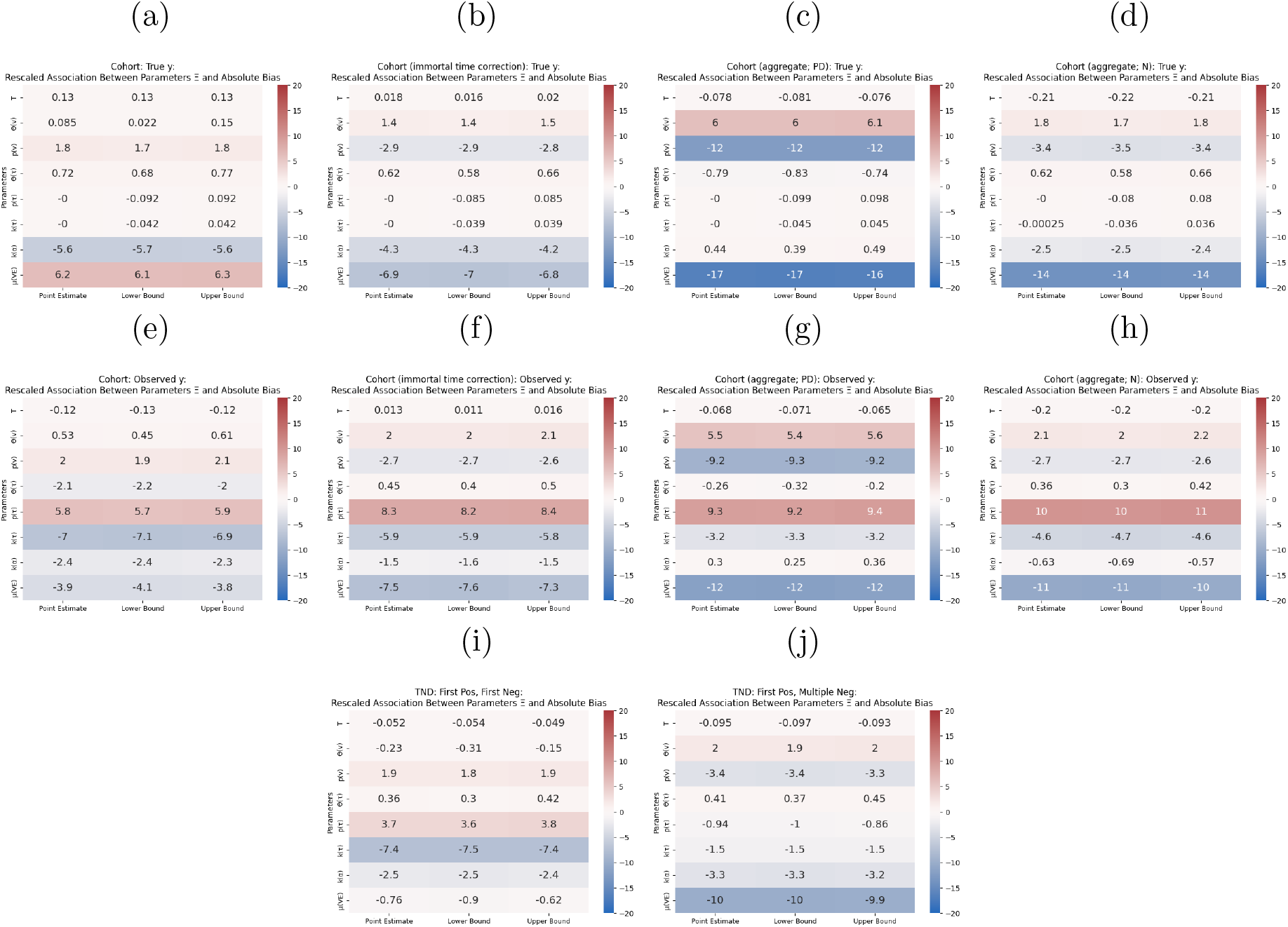
Rescaled Association Between Parameters Ξ and Absolute Bias By Study Design (*p*_*v*_ = 0.5; Θ_*τ*_ = 0.75; *p*_*τ*_ = 0.5; *k*_*τ*_ = 0.75; *k*_*α*_ = 0.75)

An alternate set of parameter values entails higher testing rate (*p*_*τ*_ = 0.75), and almost symmetric testing rate between the vax-willing and vax-unwilling (*k*_*τ*_ = 0.95). As the degree of bias arising from testing frictions almost disappeared, barring study designs whose estimation bias are highly sensitive to vaccination rates (*p*_*v*_), and asymmetry in baseline attack rates (*k*_*α*_), estimation bias can be small if confounding is absent, and if residual sources of bias are minimal, especially both TNDs. There are immense potential gains for policymakers if steps are taken to mitigate selective testing in respective infectious disease surveillance systems, such as through randomness in community sampling.

**Figure E.8:**
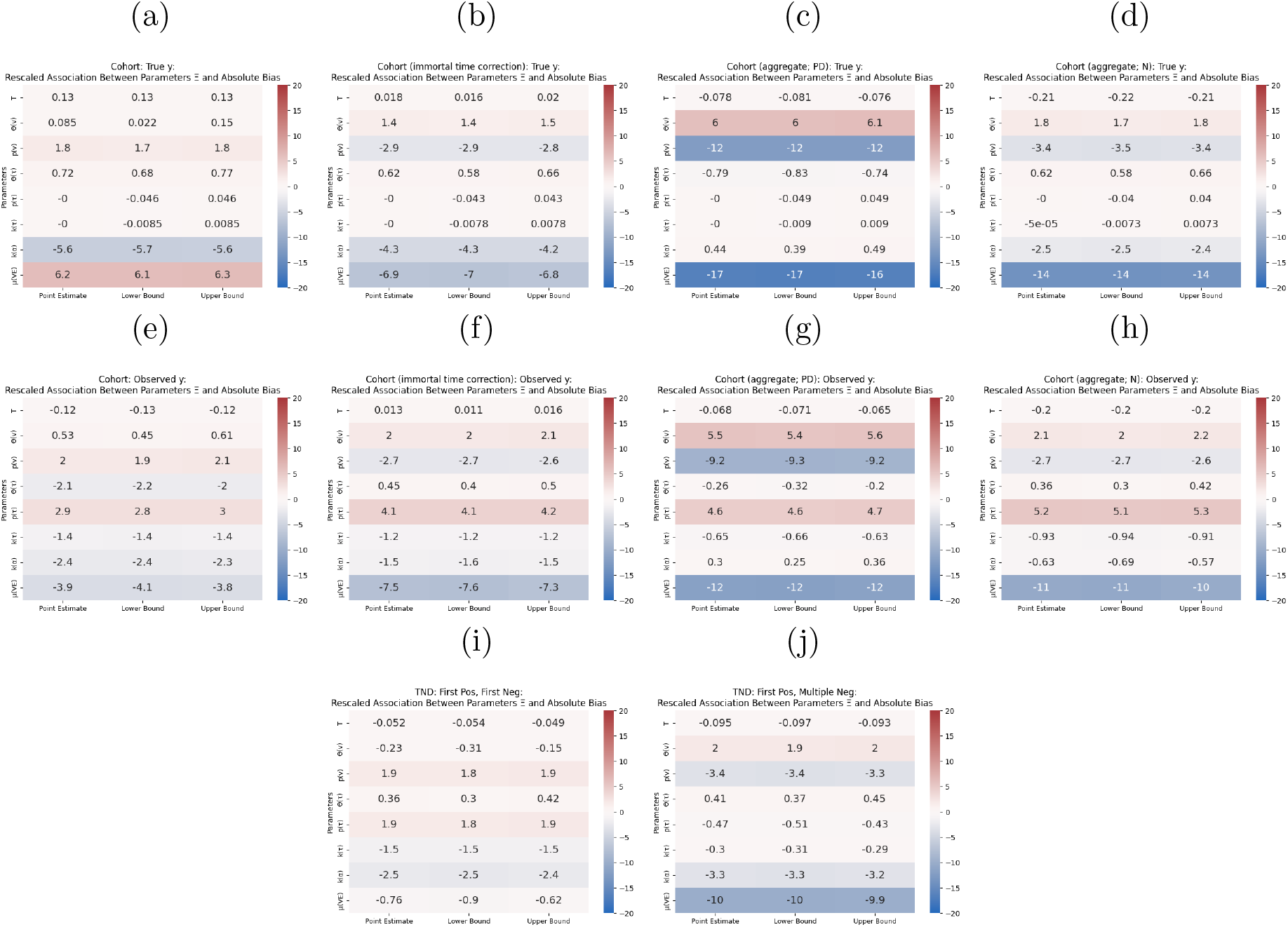
Rescaled Association Between Parameters Ξ and Absolute Bias By Study Design (*p*_*v*_ = 0.5; Θ_*τ*_ = 0.75; *p*_*τ*_ = 0.75; *k*_*τ*_ = 0.95; *k*_*α*_ = 0.75)

As a summative exercise, the FE model indicates that all factors except for follow-up duration (*T*), and deviation from full test-willing population (Θ_*τ*_), are important within-design drivers of estimation bias on average.

**Figure E.9:**
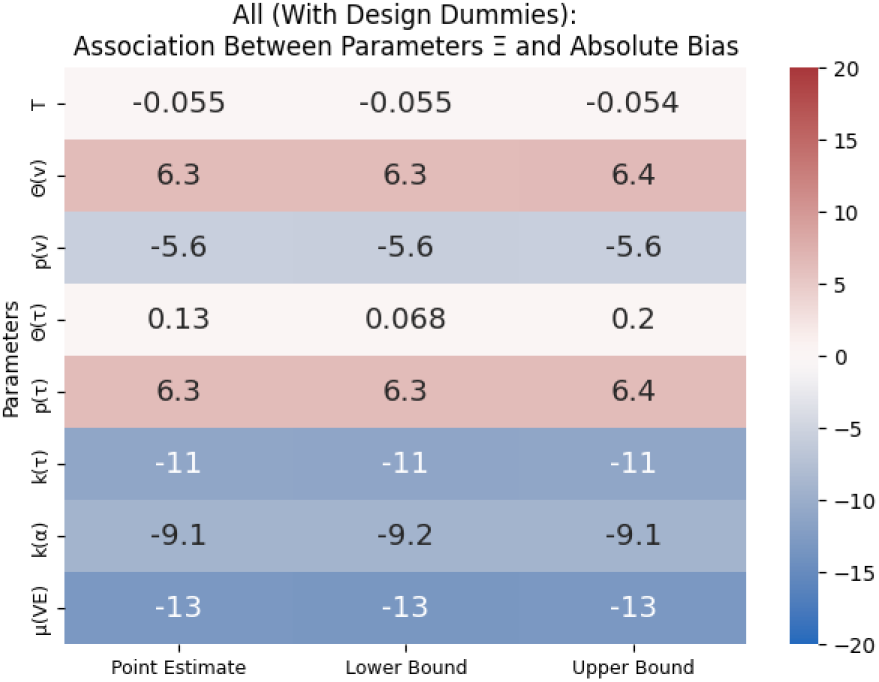
Association Between Parameters Ξ and Absolute Bias

